# Association between cannabis use and brain imaging phenotypes in UK Biobank: an observational and Mendelian randomization study

**DOI:** 10.1101/2023.08.12.23294013

**Authors:** Saba Ishrat, Daniel F. Levey, Joel Gelernter, Klaus P. Ebmeier, Anya Topiwala

## Abstract

Cannabis use during adolescence and young adulthood has been associated with brain structure and functional connectivity, yet despite a rapid increase in cannabis use among older adults in the past decade, the impact on brain structure and function in this population remains understudied. We examined 3,641 lifetime cannabis users (mean age = 61.00 years, standard deviation (SD) = 7.07) and 12,255 controls (mean age = 64.49 years, SD = 7.51) from the UK Biobank. Insufficient data were available on cannabis use disorders in the UK Biobank to perform an analysis. Brain structure and functional connectivity were measured using multiple imaging-derived phenotypes. Associations with cannabis use were assessed using multiple linear regression while controlling for potential confounders. Additionally, we conducted a two-sample Mendelian randomization analysis to investigate the potential causal relationship. After correcting for false discovery rate for multiple testing, participants with lifetime cannabis use had significantly lower fractional anisotropy and higher mean diffusivity in the genu of the corpus callosum. A weaker resting-state functional connectivity was observed in brain regions underlying the default mode and central executive networks. Furthermore, bidirectional two-sample Mendelian randomization analyses found no support for a causal relationship between either cannabis use or cannabis dependence and brain structure or function. Our findings indicate that associations between lifetime cannabis use and later life brain structure and function are not likely causal in nature and may represent residual confounding.

## Introduction

In the past decade cannabis use has increased worldwide following its legalization for medical and recreational purposes. This legalization has occurred without a comprehensive understanding of the potential effect of cannabis on the brain. Between 2006 and 2013, there was a 250% increase in reported past-year cannabis use among adults aged 65 and older in the United States.^1,2^ While cannabis use has increased in older adults, studies on health-related outcomes in this group are still limited.^3^ There are reports of adverse cannabis effects on neuro-cognitive performance, brain structure and function.^4,5^ Whether there is a safe threshold of cannabis use is unknown.

Endogenous cannabinoids, which are metabolic enzymes produced within the body, play a crucial role in various brain functions such as cognition, memory, reward processing, mood regulation, and stress sensitivity.^6^ One way in which tetrahydrocannabinol (THC), the primary psychoactive compound found in cannabis, influences the brain’s resting-state functional connectivity (rsFC) is by interacting with cannabinoid receptor type 1 (CB1). This interaction can disrupt the signaling of these naturally occurring endogenous cannabinoids, potentially affecting various brain functions.^7,8^ These acute effects of THC on the brain may be associated with chronic changes that can be detected in past cannabis users. Such effects are likely to be greater with the increased concentrations of THC found in cannabis sold after the legalization of cannabis in different parts of the world.^9^ Other cannabis harms relate to smoking, the usual method of administration, and may be attributable to other psychoactive substances than THC in cannabis.

Past use of cannabis has been linked with multiple aspects of brain structure and function in adult and adolescent populations.^10^ The most consistent brain regions linked to cannabis use are the subcortical regions. Smaller hippocampal and amygdala grey matter volume with cannabis use has been observed with long-term heavy cannabis use^11^ as well as with recent cannabis use.^12^ Diffusion tensor imaging (DTI) studies have also reported associations with white matter microstructure in the corpus callosum and forceps minor, inferred by a lower fractional anisotropy (FA) and a higher mean diffusivity (MD).^13,14^ Functional magnetic resonance imaging (MRI) studies have observed differences in functional connectivity in regions underlying default mode network (DMN) and central executive network (CEN).^15,16,17,18,19^ Whether similar brain aspects are affected in mid- to late-life adults is unclear, as only a few studies have included these age groups,^20,21^ and the focus has been on heavy or dependent users who appear to show abnormalities in structural and functional connectivity in different brain regions.^11,15,22,23^ Further, most studies have limited their analysis to a narrow range of brain regions and networks. These observational studies have been unable to distinguish between causal or confounded relationships. One of the key strengths of our study is the application of Mendelian randomization, a quasi-experimental method that uses genetic data to investigate causality.

Here, we investigate associations between cannabis use and a rich set of measures of structure and function across the brain in a large cohort of older adults. We employ both hypothesis-driven and agnostic approaches, and triangulate our observational findings with Mendelian randomization.^24^

## Methods

### Study sample

The study comprised participants from the UK Biobank listed in Supplementary material (Figure S1). For the total sample of approximately 500,000 participants, the phenotypic data used in this study were from the first repeat assessment visit (2012-2013), and the first imaging visit (2014-2019). Self-reported cannabis use data (described below) from the assessment visit were available for 157,316 participants, and MRI data from the imaging visit for 47,920 participants. Data on sociodemographic measures used in the study were collected during the imaging visit. Participants provided informed consent via electronic signature at the time of recruitment. The ethical approval for UK Biobank has been granted by the National Information Governance Board for Health and Social Care and the NHS North-West Multi-centre Research Ethics Committee.^25^

### MRI acquisition and data processing

The imaging data were obtained on Siemens Skyra 3T MRI scanners equipped with 32-channel head coils. The UK Biobank team performed image processing, quality control checks, and automated brain tissue volume computations and their imaging-derived phenotypes (IDPs) were made available to the researchers.^26^ The brain imaging protocol utilized in the UK includes structural, diffusion, and functional imaging from six distinct modalities: T1-weighted, T2-weighted flair, diffusion MRI (dMRI), susceptibility-weighted imaging (SWI), task functional MRI timeseries data (tfMRI), and resting-state functional MRI timeseries data (rsfMRI).^26, 27^

T1-weighted MPRAGE and T2-weighted FLAIR volumes were obtained at 1×1×1mm (208×256×256 field of view [FOV] matrix) and 1.05×1×1mm (192×256×256 FOV matrix) respectively. Estimation of the grey matter was performed for a total of 139 regions of interest (ROIs) defined by the HarvardOxford cortical and subcortical atlases and the Diedrichsen cerebellar atlas. Subcortical volume estimation was done using the population priors on shape and intensity variation across subjects. Image segmentation was performed to identify white matter hyperintensities (WMH). Additionally, periventricular WMH (pWMH) and deep WMH (dWMH), were defined based on subsets of total WM hyperintensities.

DMRI were obtained at 2×2×2mm (104×104×72 FOV matrix) using a multishell approach with two b-values (b = 1000 and 2000 s/mm2). 50 diffusion encoding directions were acquired for each diffusion-weighted shell and the tensor fitting was performed using the b = 1000 s/mm2. The diffusion tensor and NODDI models were applied to pre-processed data, generating nine microstructural maps: fractional anisotropy (FA), mean diffusivity (MD), axial diffusivity (L1), radial diffusivities (L2, L3) and mode of anisotropy (MO) from DTI, and intracellular volume fraction (ICVF), isotropic volume fraction (ISOVF), and orientation dispersion (OD) from NODDI. Tract-Based Spatial Statistics (TBSS) processing was performed using these DTI maps and TBSS-derived measures were computed by averaging the skeletonized images of each DTI map within a predefined set of 48 standard-space tract masks defined by the JHU White Matter Atlas (ICBM-DTI-81).^28^

SWI was obtained at 0.8×0.8×3mm (256×288×48 FOV matrix). T2* and Quantitative susceptibility mapping (QSM) were used to generate the IDPs. First T1-weighted structural brain scan was used to derive subject-specific masks for 14 subcortical regions that correspond to the left and right of the 7 subcortical structure ROIs. Subsequently, image-derived phenotypes (IDPs) were calculated based on the median T2* and χ values for each of these regions.

TfMRI were obtained at 2.4×2.4×2.4mm (88×88×64 FOV matrix) involving Hariri faces/shapes “emotion” task with either angry or fearful faces. Participants were presented with blocks of trials where they were required to determine which of the two faces displayed at the bottom of the screen correspond to the face shown at the top, or which of the two shapes presented at the bottom match the shape displayed at the top.

RsfMRI were also obtained as per tfMRI at 2.4×2.4×2.4mm (88×88×64 FOV matrix). On a preprocessed sample of 4162 participants, grouped average independent component analysis (ICA) was carried out using MELODIC.^27, 29^ ICA was performed with dimensionality set to 25 and 100, which resulted in 21 and 55 components, respectively, after discarding the noise components. These 21×21 and 55×55 partial correlation matrices were used as measurements of functional connections. The ICA maps were mapped on each participant’s rsfMRI timeseries data in order to acquire one representative node timeseries per ICA component for each subject. These network ‘nodes’ are a measure of within-network functional connectivity. Subject-specific network-matrices (‘edges’) were also extracted from the node timeseries that provide a measure of functional connectivity between the nodes.^30^

### Cannabis use data

Cannabis use was self-reported at the online follow-up during the first repeat assessment visit. Participants reported if they had “Ever taken Cannabis”. Possible answers were: ‘no’, ‘prefer not to say’, ‘yes, 1–2 times’, ‘yes, 3–10 times’, ‘yes, 11–100 times’, and ‘yes, more than 100 times’ **(SFigure 1).** All participants who responded ‘yes’ were categorized as lifetime cannabis users, and ‘no’ responders were categorized as controls. Cannabis users were further divided into two subgroups: (a) low-frequency cannabis use (lifetime cannabis use of 1-10 times), and (b) high-frequency cannabis use (lifetime cannabis use of 11-100+ times). This subgroup categorization for cannabis users was introduced in a previous study.^31^ Participants also reported their “Age when last taken cannabis” and we computed years since the participants last had cannabis by the difference between the age when last cannabis was used and the age when subjects were scanned. Additionally, only two participants were identified with cannabis use disorder in UK Biobank, which was insufficient for conducting an analysis.

### Genetic variants

We examined summary genomewide data based on two different cannabis phenotypes. Detailed information on the single nucleotide polymorphisms (SNPs) used to instrument these phenotypes is provided in **STable 1**.

For the first exposure variable, we used the genome-wide association study (GWAS) summary statistics for individuals of European ancestry on *cannabis dependence or abuse* consisting of 15 cohorts. Ancestry-specific linkage disequilibrium clumping was performed using PLINK v2.0 with the respective 1000 Genomes Project phase 3 linkage disequilibrium reference panels. Lead variants were identified within 10,000 kb and LD r^2^=0.001. This GWAS identified 23 genome-wide significant independent SNPs. The IDPs used as our outcome variable had 20 matching SNPs, so we identified proxy SNPs (R^2^C>C0.9) from LDlink (https://ldlink.nci.nih.gov/). Of the missing 3 SNPs, one SNP was monoallelic leaving a total of 22 SNPs in our analysis (**STable 1 (a)**).^32^

The second exposure variable consisted of the GWAS summary statistics for *lifetime cannabis use* from the International Cannabis Consortium (ICC), 23andMe and UK Biobank (N = 184,765) from individuals of European ancestry. Participants reported if they had ever used cannabis during their lifetime and the response was recorded as yes or no. This GWAS identified 8 genome-wide significant independent SNPs. The estimated SNP-heritability (h^2^_SNP_) for *lifetime cannabis use* was 11% (**STable 1 (a**)).^33^

We obtained the summary statistics for each of the brain IDPs as the outcome variable from the GWAS performed by the UK Biobank, which included approximately 33,000 participants.^34^

For the reverse MR analysis, the brain IDPs GWAS identified 1 SNP associated at genome-wide significance (p<5×10^−8^) with the FA of the genu of the corpus callosum, 2 SNPs associated with rfMRI connectivity (ICA25 edge 21 and ICA100 edge 55) that matched with SNPs in *cannabis dependence or abuse* GWAS. Notably, only one SNP associated with rfMRI connectivity (ICA25 edge 21) had a matching SNP in *lifetime cannabis use* GWAS (**STable 1 (b**)).

### Confounds

We adjusted for potential confounds, which were self-reported *at the time of the MRI scan*. Age at first scan (in years), sex (male and female), and also age^2, age^3, and age-by-sex interaction were controlled for. Townsend deprivation is a measure of material deprivation based on census information. Current employment status was recorded as: in paid employment/self-employed, retired, looking after home and/or family, unable to work because of sickness or disability, unemployed, doing unpaid or voluntary work, full or part-time student or none of the above. Educational qualifications were recorded as: college or university degree, A level/AS levels or equivalent, O levels/GCSEs or equivalent, CSEs or equivalent, NVQ or HND or HNC or equivalent, other professional qualifications or none of the above. Smoking and alcohol drinking status was reported as: current, previous or never. Systolic and diastolic blood pressure (DBP) were measured in mmHg and body mass index (BMI) in kg/m^2^. For a measurement of mental health status, participants were asked if they had ‘seen a psychiatrist for nerves, anxiety, tension or depression’ and the response was noted as ‘yes’, or ‘no’.

We also accounted for a set of 613 brain imaging-related confounds in this sample as described in Alfaro-Almagro et al (2020).^35^ These included: assessment centre, intracranial volume, head motion, table position, and scanner acquisition parameters (site, scanner software, protocol, scan ramp, head coil).

### Statistical analyses

All statistical analysis was performed in R (version 4.0.0) and visualizations were performed in MATLAB (version R2018_a). Independent samples *t*-tests and chi-squared analyses were performed to assess potential univariate differences in the sociodemographic characteristics between the cannabis users and controls. Multiple linear regression was performed to determine the relationship between cannabis use and brain measures, accounting for confounds.

To begin with, in a hypothesis-driven approach, we examined the association between cannabis use and grey matter volume of the hippocampus and amygdala to test the hypotheses from previous studies that were conducted on samples consisting of adolescents and young adults. Subsequently, we employed an exploratory approach to examine the association between cannabis use and brain structure and function by utilizing all brain IDPs. A total of 3,921 brain IDPs were tested with an adjusted cut-off p-value of 0.05 using Bonferroni correction. For comparison, p-values in the analyses were also corrected for multiple testing using false discovery rate (FDR, 5%).

We then performed sensitivity analyses controlling for the covariates. These were performed for the cannabis-IDPs associations that remained statistically significant after adjusting for multiple comparisons using the FDR test. We performed two sensitivity analyses amongst cannabis users to assess whether: 1) years of cannabis abstinence, and 2) cannabis dose (low vs. high frequency), modified cannabis-brain associations.

Finally, we performed two-sample MR analyses by using the *TwoSampleMR* in an R package^36^ to investigate whether significant observed associations between cannabis use and brain IDPs were causal. P-values in the analyses were additionally adjusted for multiple testing FDR. For the SNPs significantly associated with *cannabis dependence or abuse* and *lifetime cannabis use,* five different MR methods were applied: inverse-variance weighted (IVW), MR Egger, weighted median, weighted mode, and simple mode. The various MR methods make distinct assumptions regarding the nature of pleiotropy. The IVW method was utilized because the instruments used in the study consisted of multiple SNPs. To ensure valid results, the IVW method requires that all instruments are associated with the exposure variable, but not directly associated with the outcome variable or any confounding factors affecting the relationship between the exposure and outcome. MR Egger tests for horizontal pleiotropy, which assumes that the pleiotropy effects are independent of the strength of the instrument used. The estimate obtained from the weighted median gives an estimate of the causal effect when at least half of the genetic variants used are valid instrumental variables. Weighted mode assumes that the validity of the instruments is based on the largest number of instruments with consistent MR estimates. Last, the simple mode approach uses the mode of the instrumental variable estimate to estimate the causal effect. In addition, we performed a reverse analysis to test for reverse causation.

## Results

### Demographics

There were 3,641 cannabis users and 12,255 control subjects with complete data (**SFigure 2**). Only two individuals had cannabis abuse or dependence codes in the linked electronic health record. Cannabis users were significantly younger than non-users (**Table 1**). While the subjects were well matched for BMI and diastolic BP, the user group had significantly lower systolic BP and were less socially deprived than the control group. There was a slightly higher proportion of males in the user group, a higher proportion of the user group were in employment and had college degrees. A higher proportion of cannabis users than controls drank alcohol and smoked cigarettes. Additionally, a higher proportion of the user group complained of nerves/anxiety/tension/depression (**Table 1**).

**Table 1:** Demographic characteristics. Abbreviations: SD, standard deviation; BMI, body mass index; BP, blood pressure

| Variables (at time of imaging) | Cannabis users (n = 3,641) | Controls (n = 12,255) | Statistics |  |
| --- | --- | --- | --- | --- |
|  | *Mean (SD) or % (n)* |  | *t or $\chi^2$\** | *p-value* |
| Age at 1<sup>st</sup> scan (years) | 61.00 (7.07) | 64.49 (7.51) | 24.94 | 2.20e-16 |
| Sex (male % (n)) | 52.40% (1,908) | 43.96% (5,387) | 80.29* | 2.20e-16 |
| Townsend deprivation index | -1.23 (2.99) | -2.24 (2.50) | -20.31 | 2.20e-16 |
| Employment status (current; % (n)) | 52.89% (1,962) | 34.30% (4,204) | 519.4* | 2.20e-16 |
| College degree % (n)) | 65.42% (2,382) | 46.37% (5,683) | 477.89* | 2.20e-16 |
| BMI (kg/m<sup>2</sup>) | 26.21 (4.23) | 26.37 (4.36) | 1.97 | 0.05 |
| Diastolic BP (mmHg) | 78.54 (10.46) | 78.69 (10.70) | 0.76 | 0.45 |
| Systolic BP (mmHg) | 137.36 (19.18) | 141.19 (19.88) | 10.30 | 2.20e-16 |
| Alcohol use (current; % (n)) | 96.26% (3,505) | 93.43% (11,450) | 93.47* | 2.20e-16 |
| Smoking (current; % (n)) | 7.39% (269) | 1.93% (337) | 1320.10* | 2.20e-16 |
| Nerves, anxiety, tension, or depression status (% (n)) | 11.40% (415) | 7.90% (968) | 42.83* | 5.98e-11 |

### Observational Analysis

In view of previously observed associations with hippocampal and amygdala volume, we examined these as regions of interest in a hypothesis-driven approach. There were no significant associations observed with cannabis use (**STable 2**).

Out of 3,921 brain IDPs, cannabis use was significantly associated with 40 brain IDPs after *FDR* correction (0.05%, p = 0.009) (**Figure 1**, **Table 2**, **STable 3**). The strongest associations were with measures of white matter microstructure. Most significant associations identified in the DTI metrics were found in the genu and body of the corpus callosum, demonstrating lower fractional anisotropy (FA) and intracellular volume fraction (ICVF), as well as higher mean diffusivity (MD), and radial diffusivity (L2, L3). Furthermore, a higher MD was observed in the left cingulum cingulate gyrus, while increased L2 was detected in the cingulum bundle, and higher L2 and L3 were observed in the anterior corona radiate (**Figure 2**).

**Figure 1:**
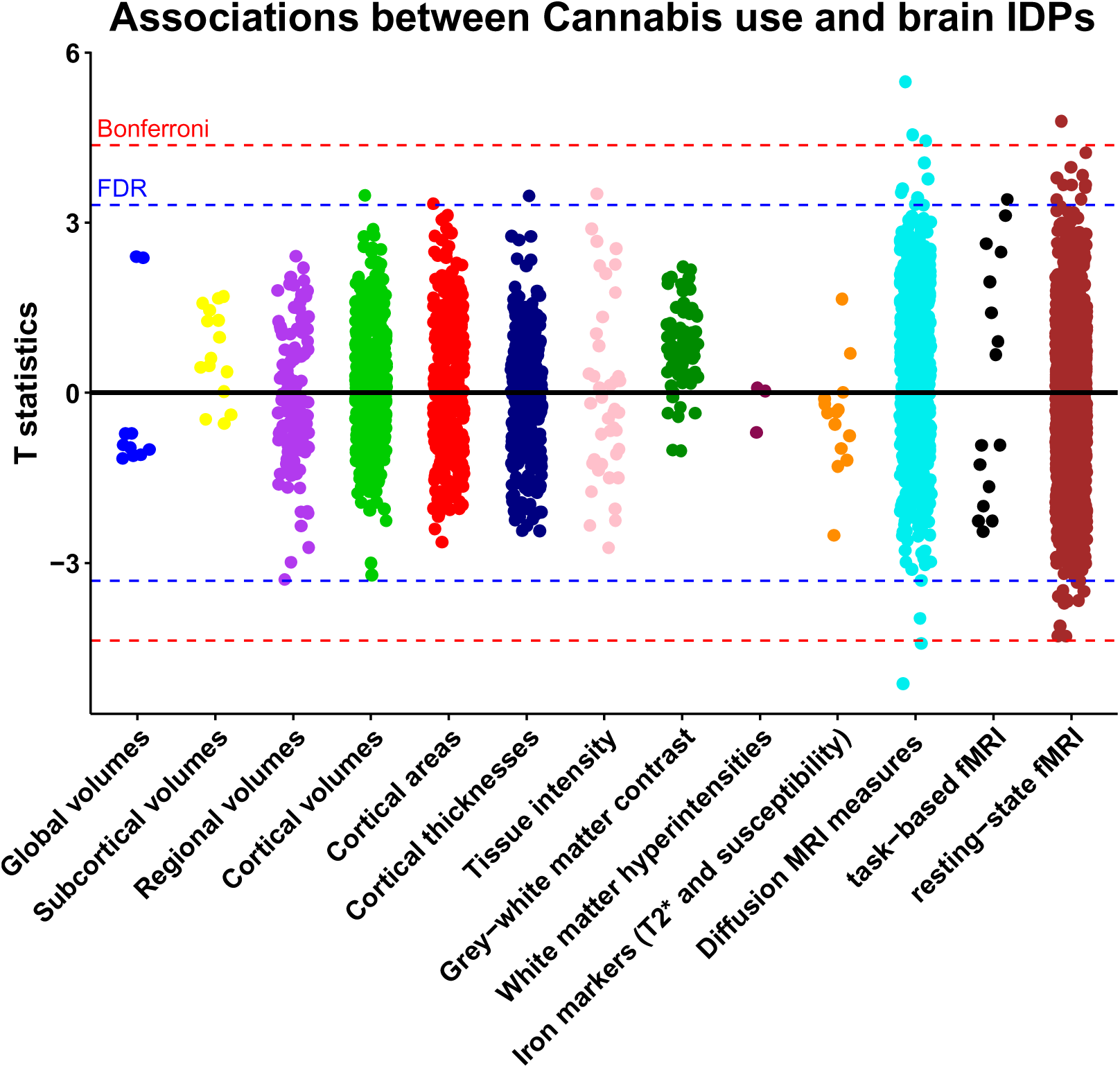
Associations between cannabis use and brain image-derived phenotypes. Estimates were generated using multiple linear regression models adjusted for: age, sex, Townsend deprivation index, employment status, educational qualifications, alcohol drinking status, smoking status, body mass index, systolic and diastolic blood pressure, assessment center, nerves/anxiety/tension/depression status and brain imaging confounds. Red line indicates the Bonferroni threshold (3,921 tests, p = 1.28 × 10^−5^, T statistics = 4.36) and blue line indicated the False Discovery rate threshold (3,921 tests, p = 9.38 × 10^−4^, T statistics = 3.31). Abbreviations: MRI, magnetic resonance imaging; fMRI, functional magnetic resonance imaging

**Figure 2:**
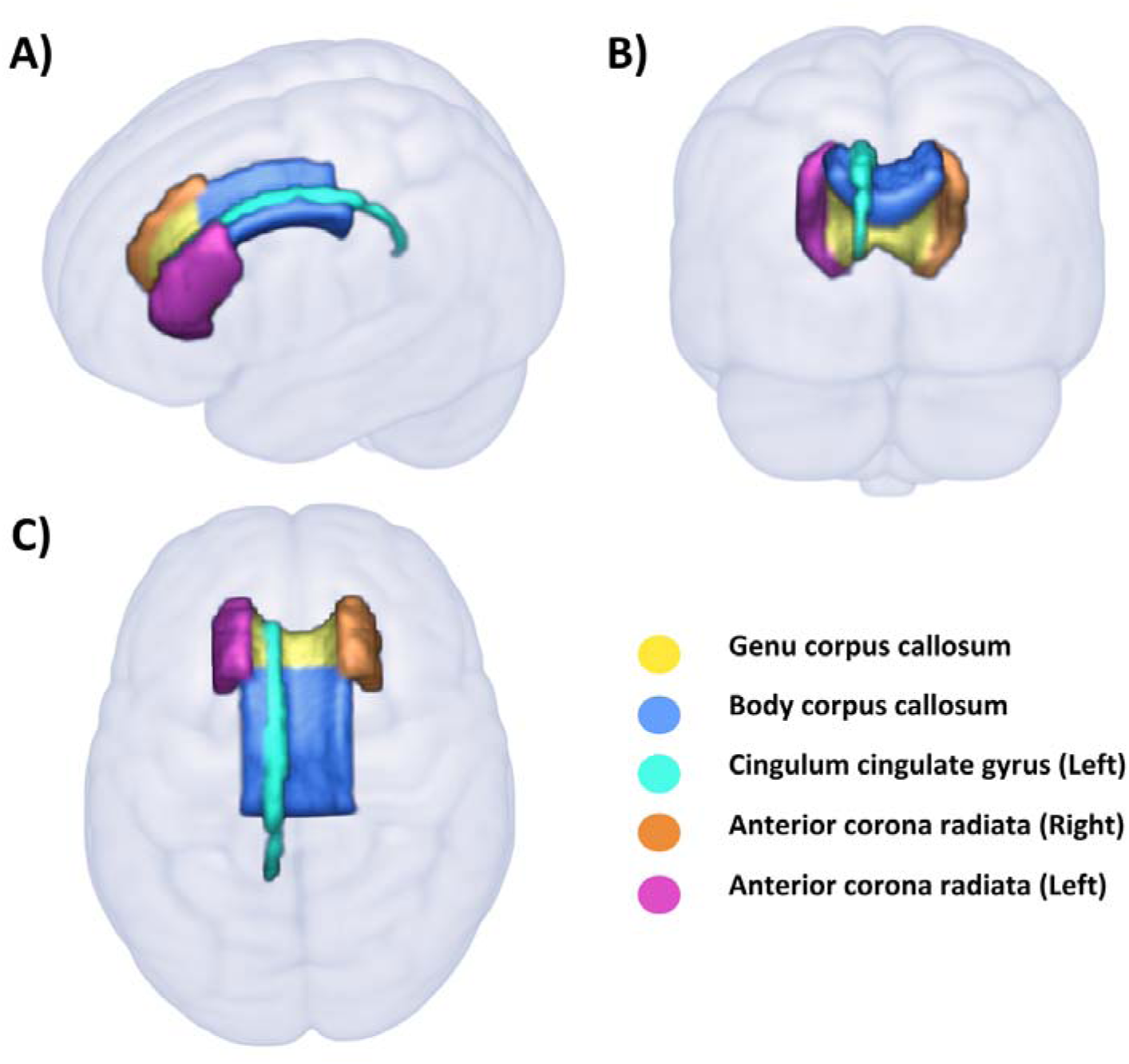
White matter regions significantly associated with cannabis use. Structures delineated by the JHU White Matter Atlas (ICBM-DTI-81) presented in A) sagittal, B) coronal, and C) axial views.

**Table 2:** Summary of the association between cannabis use and brain measures after False Discovery Rate correction (5%). Abbreviations: MRI – magnetic resonance imaging; FA, fractional anisotropy; ICVF intracellular volume fraction; MD, mean diffusivity; L2 and L3, radial diffusivities.

| Brain measures | Regions/Tasks/Networks |  | Direction/strength of association with cannabis use |
| --- | --- | --- | --- |
| Volume | Right inferior lateral ventricles |  | Positive |
| Area | Left frontal pole |  | Positive |
| Thickness | Left posterior ventral cingulum gyrus |  | Positive |
| Regional and tissue Intensity | Right pallidum |  | Positive |
| Diffusion tensor imaging | FA | Genu and body of corpus callosum | Negative |
|  | ICVF | Genu of corpus callosum |  |
|  | MD | Genu of corpus callosum, cingulum cingulate gyrus | Positive |
|  | L2 | Genu and body of corpus callosum, and left cingulum cingulate gyrus |  |
|  | L3 | Genu and body of corpus callosum, genu, and right and left anterior corona radiata |  |
| Task-based functional MRI | Faces-shapes 'Emotion' task |  | Positive |
| Resting state functional MRI | Default mode network, central executive network, salience network, and motor network, |  | Stronger |
|  | Default mode network, central executive network, salience network, motor network, visual network, subcortical-cerebellum network, attention network and limbic network |  | Weaker |

A wide range of associations was observed across various rsFC analyses, particularly indicating either weaker or stronger connectivity between multiple networks. These networks predominantly included brain regions associated with the Default mode, Central executive, and Salience network. A visual representation of the resting-state networks as nodes and their connections that are significantly associated with cannabis use for both 21 and 55 resting-state networks obtained, respectively, from 25-component and 100-component group-ICA, is presented in **Figure 3 and STable 4**.

**Figure 3:**
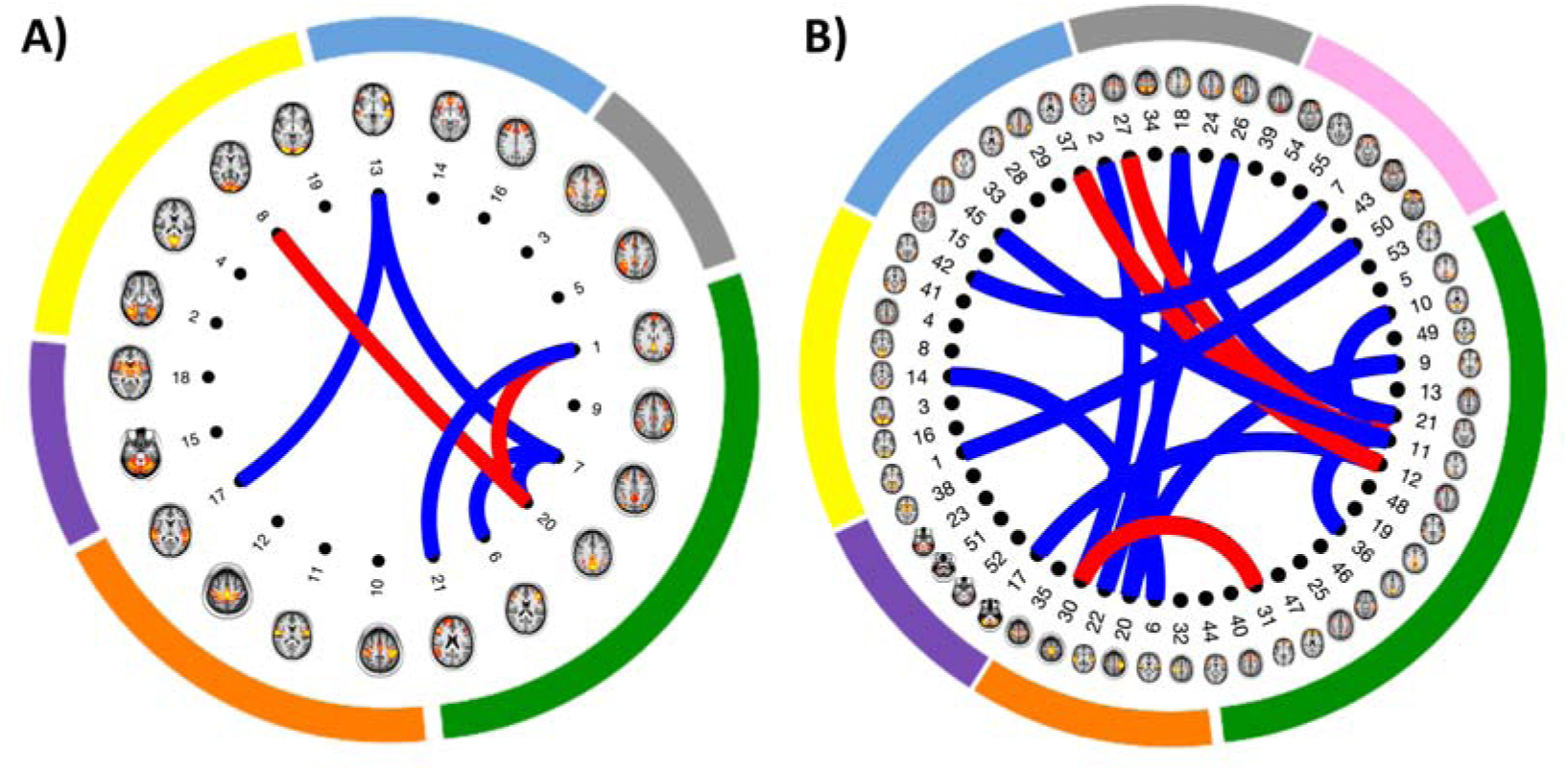
Resting state functional connectivity significantly associated with cannabis use. Spatial maps of resting state networks nodes (n=21 and n=55) are illustrated in brain images. These were identified as non-noise components from: A) 25-component group-independent component analysis proposed by Miller et al 2016.^26^ Nodes are grouped by networks as follows: default mode/central executive network (green), motor network (orange) subcortical-cerebellum network (purple), visual network (yellow), salience/default mode/central executive network (blue) and attention/default mode/central executive network (grey). B) 100-component group-independent component analysis. Nodes are grouped by networks as follows: default mode/central executive network (green), motor/attention network (orange) subcortical-cerebellum network (purple), visual/attention network (yellow), salience/default mode/central executive network (blue), attention/salience/central executive network (grey) and limbic/default mode network (pink). Connecting lines indicate partial correlations between network nodes significant associated with cannabis (red lines indicate stronger connectivity and blue lines indicate weaker connectivity).

Cannabis use also associated with specific brain IDPs including a larger volume of the right inferior lateral ventricles, a larger surface area of the frontal pole, greater thickness in the posterior ventral cingulum gyrus, higher intensity in the right pallidum, and enhanced group-average BOLD activation to emotional faces/shapes during tfMRI.

Associations with six brain IDPs additionally survived the more stringent *Bonferroni-corrected* threshold (p =1.275 × 10^−5^). Cannabis users were characterized by lower FA and ICVF, and higher MD, L2, and L3 in the genu of the corpus callosum. Additionally, cannabis users revealed lower functional connectivity between brain regions in the inferior frontal, middle frontal, and precuneus regions, all of which are associated with the default mode and central executive network (**Table 3**).

**Table 3:**
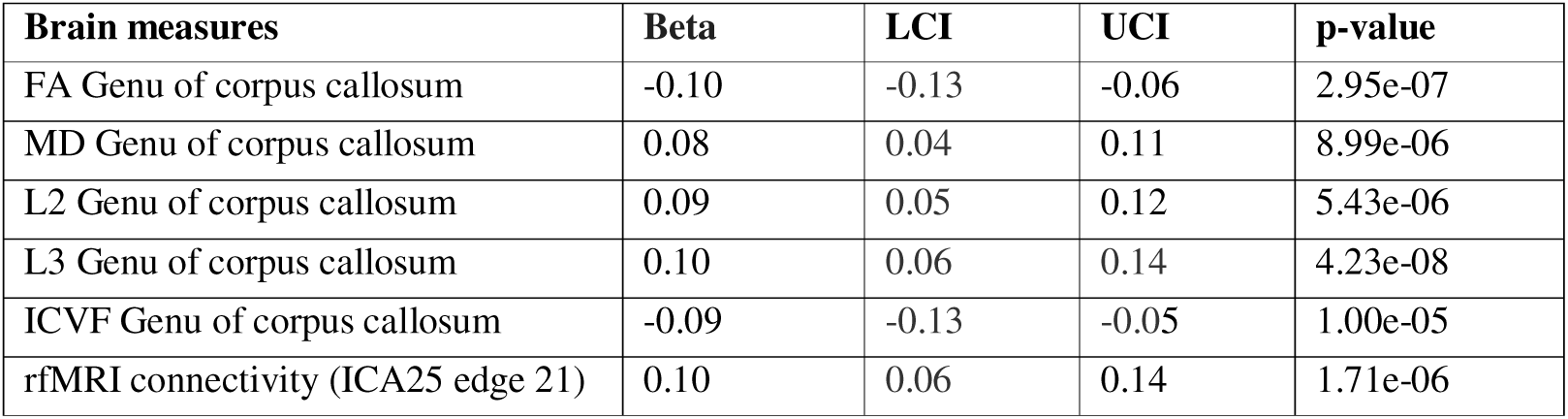
Associations between cannabis use and brain measures surviving Bonferroni correction. Estimates represent beta coefficients from multiple linear regression models adjusted for: age, sex, Townsend deprivation index, employment status, educational qualifications, alcohol drinking status, smoking status, body mass index, systolic and diastolic blood pressure, assessment centre, nerves/anxiety/tension/depression status and brain imaging confounds. Abbreviations: LCI, lower confidence interval; UCI, upper confidence interval; FA, fractional anisotropy; MD, mean diffusivity; L2 and L3, radial diffusivities; ICVF intracellular volume fraction; rfMRI, resting-state functional magnetic resonance imaging.

### Sensitivity Analyses

We assessed whether the duration of abstinence or dose impacted the relationships between cannabis use and brain IDPs that survived the false discovery rate correction in the main analysis. Neither the duration of cannabis abstinence nor frequency of cannabis dosage (as assessed through low and high-frequency use) significantly moderated the associations between cannabis use and brain measures.

### Mendelian randomization

There were no significant associations between either genetically predicted *cannabis dependence or abuse* (IVW beta= 0.01 [95% CI −0.09 to 0.11], p = 0.85), or *lifetime cannabis use* (IVW beta= −0.05 [95% CI −0.15 to 0.05], p = 0.33) with FA genu of corpus callosum after correcting for multiple comparisons. No significant association was observed with other brain IDPs either. There was no indication of horizontal pleiotropy as determined by the MR-Egger intercept test for any of the outcomes **(STable 5 (a) and SFigure 3 (a))**. Reverse MR analysis did not show any significant association between brain IDPs and *cannabis dependence or abuse* or *lifetime cannabis use* (**STable 5 (b) and SFigure 3 (b)**).

## Discussion

To the best of our knowledge, this is the largest observational study of relationships between cannabis use and brain structure and function to date, and the first Mendelian randomization investigation. Cannabis users had significant differences in brain structure and function, most markedly markers of lower white matter microstructure integrity. Genetic analyses found no support for causal relationships underlying these observed associations.

Cannabis users showed a lower fractional anisotropy and a higher mean diffusivity in the genu of the corpus callosum compared to non-users. Previous studies on the frequent use of high-potency cannabis by adolescents and young adults have reported disruption in the corpus callosum integrity. Take together, this suggests that the corpus callosum might be particularly sensitive to high tetrahydrocannabinol concentration.^37^ Cannabis users had lower white matter integrity, as proxied by higher radial diffusivities (L2, L3) of the anterior corona radiata and a higher mean diffusivity in the left cingulum. Although no associations in these diffusion metrics were reported or observed in past studies, disrupted microstructural integrity in these tracts was reported with other diffusion measures showing decreased fractional anisotropy in anterior corona radiate and increased fractional anisotropy of the cingulum.^38, 39^

Cannabis use significantly associated with resting-state functional connectivity mainly in the brain regions underlying default mode, central executive and salience networks. The brain regions underlying these networks were primarily located in the frontal lobe, temporal lobe, occipital cortex, supplementary motor area, precuneus, and cerebellum. These regions are characterized by a high density of cannabinoid receptor type 1, and have also been implicated in younger cannabis users.^15,22,40^ Higher functional connectivity between the prefrontal cortex and occipital cortex was reported in young adult chronic cannabis users compared to controls^15^. Altered patterns of functional connectivity, mostly a hypoconnectivity has been reported from the cerebellum to specific cortical regions including the prefrontal gyrus, disrupting the information flow that is essential for cognitive control and emotional regulation.^38^ Additionally, increased functional connectivity was reported in regions underlying orbito-frontal cortex with precuneus and cerebellar regions, which may be an indication of impairment in decision-making capacity and an increase in impulsive behaviour.^22^ Our findings however show a complex pattern of higher or lower functional connectivity between these regions with cannabis use.

There were also a few other brain structures that showed associations with cannabis use. Cannabis users showed a higher area of the left frontal pole and higher tissue intensity in the right pallidum. While the literature has not previously reported any similar associations with these measures, others have reported a higher volume and a morphological deformation of the pallidum,^41,42^ and a decreased gyrification of the frontal pole.^43^ We also observed novel associations with left inferior lateral ventricle volume and left posterior ventral cingulum gyrus thickness.

We did not replicate previously observed associations between cannabis use and grey matter volume in the hippocampus and amygdala. One possible explanation is the differing age range of subjects. Previous studies examined adolescents and young adults whereas our sample was middle- to late-life adults. White matter microstructural changes may also be more sensitive to cannabis effects than the grey matter measures used in this study. Longitudinal studies on the effect of cannabis on grey and white matter are needed to further investigate whether changes occur over time.

We found no influence between the duration of abstinence from cannabis use prior to the brain scan and structural or functional brain connectivity. Additionally, we did not find any significant differences in structural and functional connectivity between low and high-frequency cannabis users, thus indicating the absence of a dose-response relationship. This may be a result of our sample, consisting of healthy volunteers and a few high cannabis users.

Mendelian randomization provided no support for a causal effect of cannabis use or dependence on brain structure or function, nor a causal effect of brain structure or function on cannabis use. The disparity between observational and Mendelian randomization findings could result from several phenomena. First, the observational associations may be confounded by an unmeasured variable such as family history, dietary intake or use of certain medication. Second, our Mendelian randomization analyses had less statistical power than observational analyses to detect small effects. Future larger-scale neuroimaging GWAS will be helpful in distinguishing these two hypotheses. Lastly, Mendelian randomization assesses the lifelong impact of cannabis use, while changes observed in observational studies could occur at different life stages.

## Limitations

Our study has several limitations. First, although our sample size was bigger than that of previous studies, UK Biobank is healthier than the general population. It suffers from selection bias with respect to sociodemographic variables, such as physical, lifestyle and health-related characteristics ^44^ Furthermore, this sample is not suitable for examining cannabis use disorders due to the limited number of participants with cannabis use disorder. Second, as the age of cannabis initiation was not assessed, we were unable to examine the critical time points during life for cannabis effects. Third, participants are susceptible to recall and reporting bias concerning the amount or frequency of cannabis intake in their lifetime, and their response might be affected by external biases arising from social desirability. Determining historical cannabis use is challenging. For instance, urine drug screens typically have a limited detection period, usually just a few days. Fourth, the self-report did not include the potency of cannabis consumed, which might differ across the participants and over time. Fifth, despite our effort to account for potential confounds in our study, the presence of unmeasured confounding variables (residual confounding) cannot be excluded. Sixth, neuroimaging measures were cross-sectional and longitudinal relationships cannot be inferred. Finally, Mendelian randomization relies on several assumptions, some of which not testable.

## Conclusion

Lifetime cannabis use is associated with several measures of brain structure and function in later life, particularly in the corpus callosum. Genetic analysis did not provide support for these associations resulting from causal relationships, suggesting residual confounding may be responsible.

## Data Availability

All data produced are available online at:
https://biobank.ndph.ox.ac.uk/showcase/search.cgi

## Acknowledgments

This work was undertaken using UK Biobank application numbers 55929 & 8107. Saba Ishrat is supported by the Society for the Study of Addiction (Registered Charity No. 1009826) and Anya Topiwala by a Wellcome Trust Clinical Research Career Development Fellowship (216462/Z/19/Z). The Wellcome Centre for Integrative Neuroimaging is supported by core funding from the Wellcome Trust (203139/Z/16/Z). This research used data from the Million Veteran Program and was supported by funding from the Department of Veterans Affairs Office of Research and Development, Million Veteran Program Grant #I01CX001849, and the VA Cooperative Studies Program (CSP) study #575B, and MVP025. This publication does not represent the views of the Department of Veterans Affairs or the United States Government.

