## Supplementary figures and images for "Association between cannabis use and brain imaging phenotypes in UK Biobank: an observational and Mendelian randomization study"

### Supplemental Figure 1

**SFigure 1: Lifetime cannabis use in UK Biobank participants analysed.**

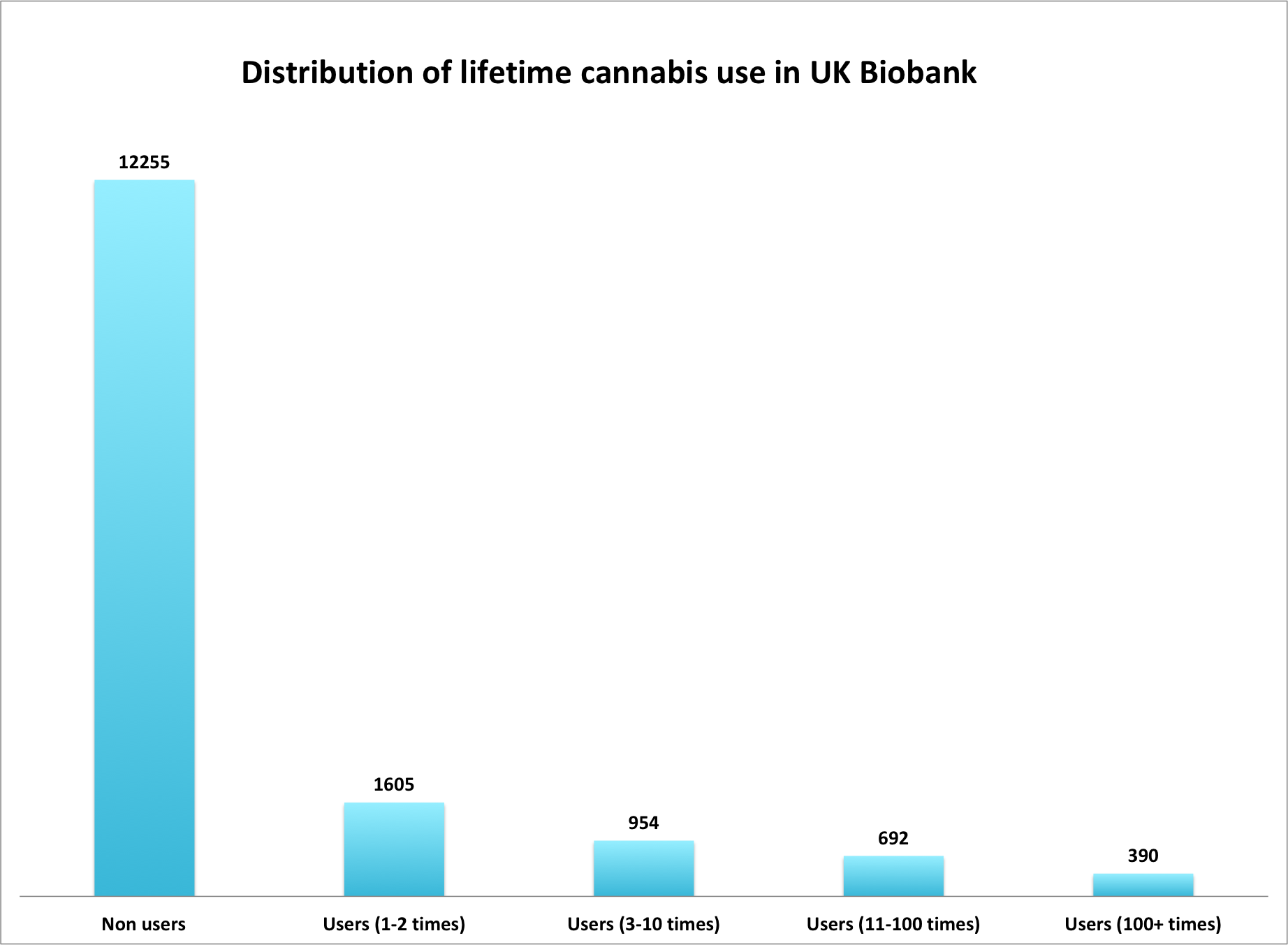
