## Supplemental Figure 2 for "Association between cannabis use and brain imaging phenotypes in UK Biobank: an observational and Mendelian randomization study"

**SFigure 2: Flow chart of the participants included in the final analysis.**

UK Biobank participants (n = 500,000)

Participants excluded with no data on cannabis (n= 342,684)

157,316 participants data on cannabis use

Participants excluded with no MRI or missing confounder data (n = 135,442)

15,896 participants with complete relevant data

Cannabis users (n = 3,641)

Controls (n = 12,255)

Low-frequency cannabis users (n = 2,036)

High-frequency cannabis users (n = 1,605)

Low-frequency cannabis use defined as lifetime cannabis use of 1-10 times, and high-frequency cannabis use defined as lifetime cannabis use of 11-100+ times.

Abbreviations: MRI, magnetic resonance imaging.
