## Supplemental Figure 3 for "Association between cannabis use and brain imaging phenotypes in UK Biobank: an observational and Mendelian randomization study"

**SFigure 3 (a)**

**Two-sample linear MR plot for the causal effect of Cannabis dependence or abuse on brain IDPs**

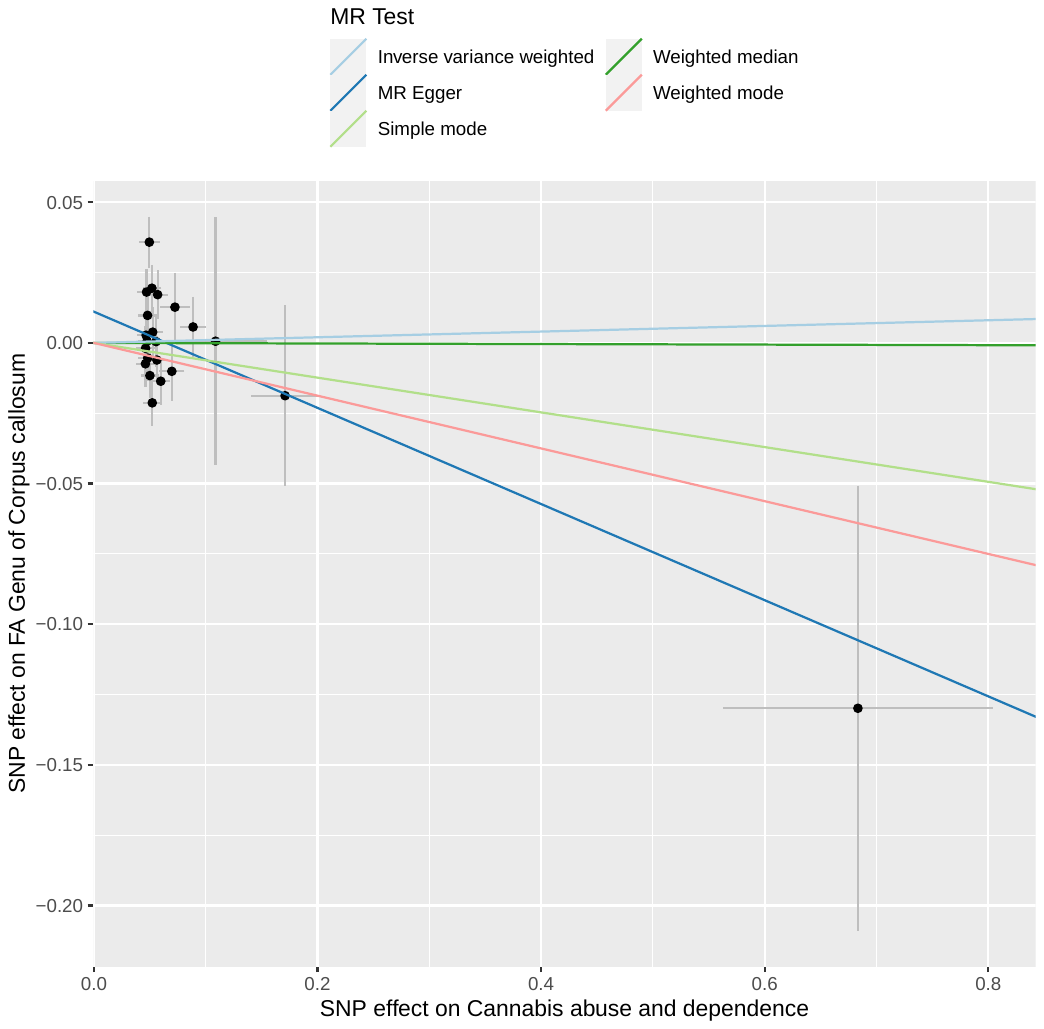

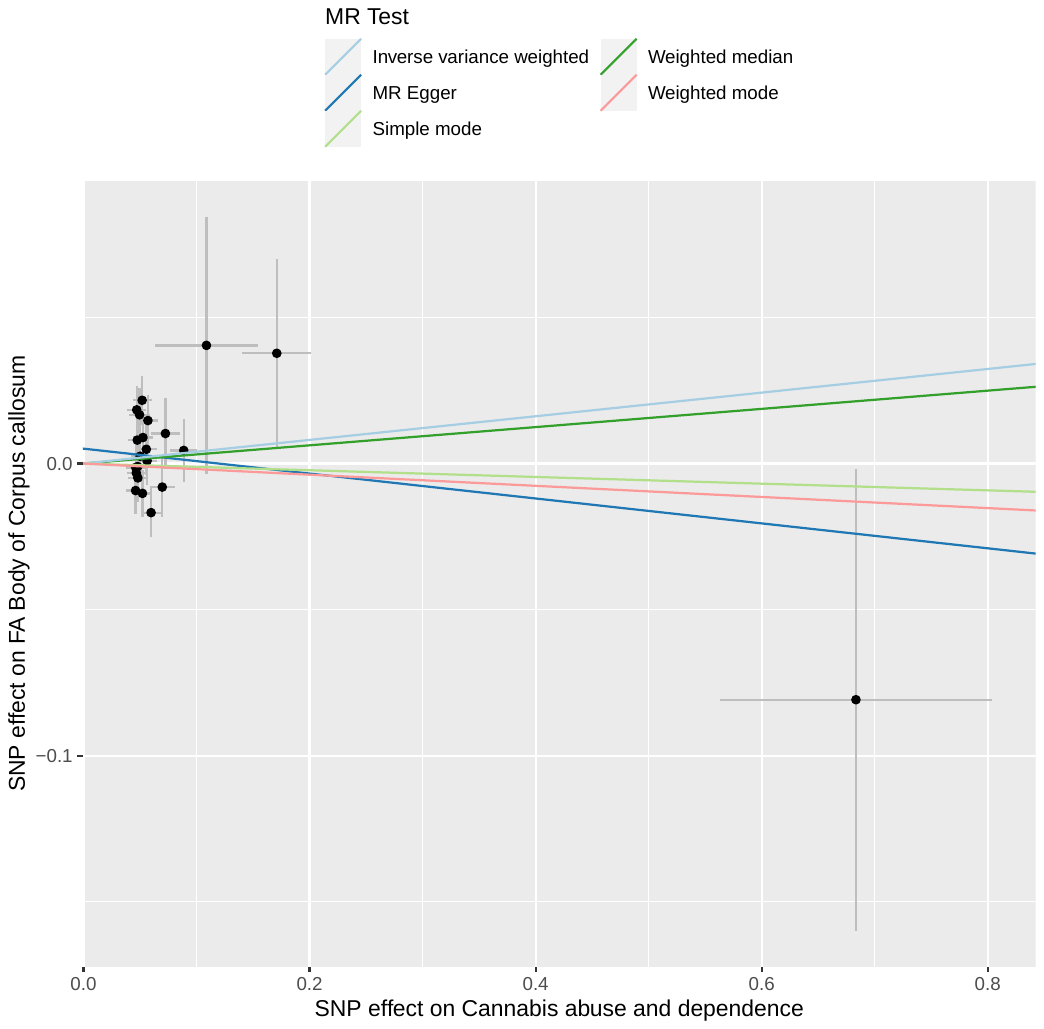

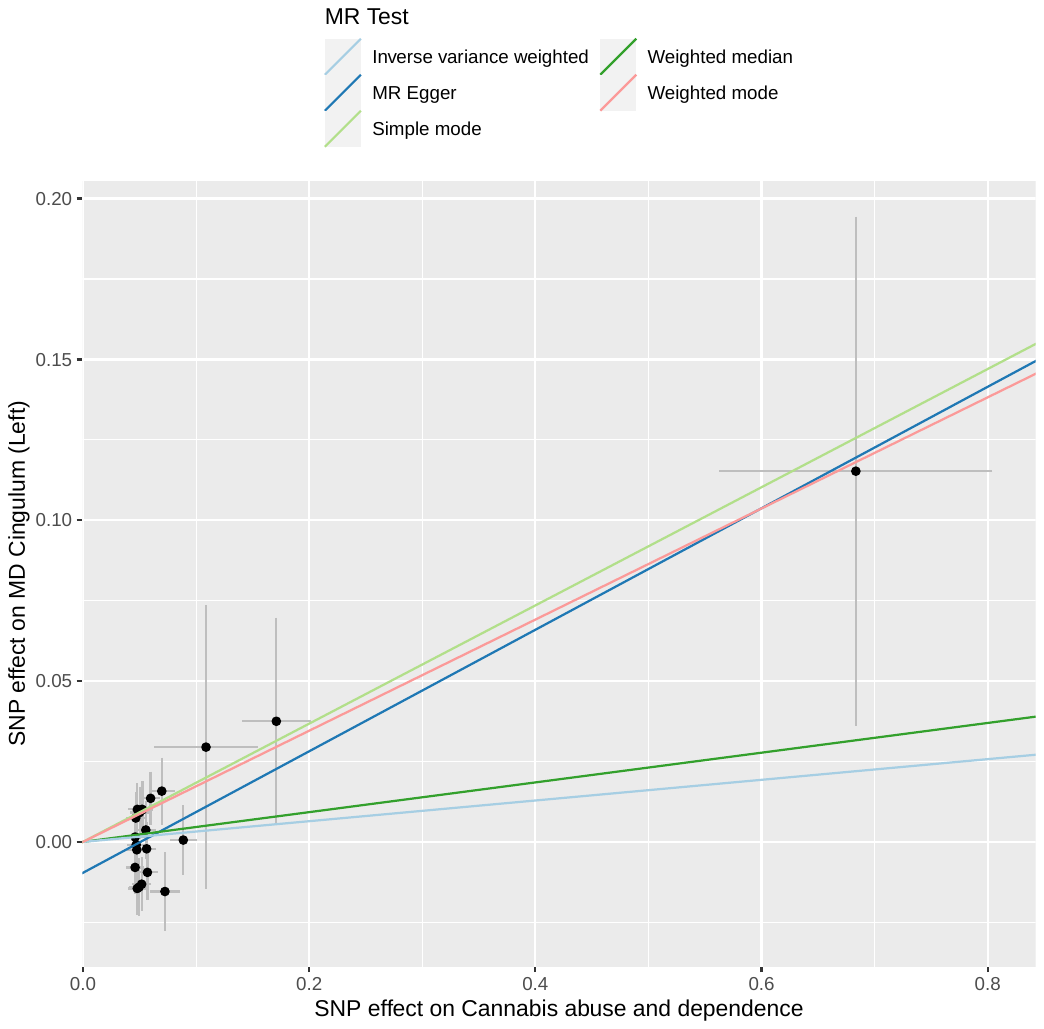

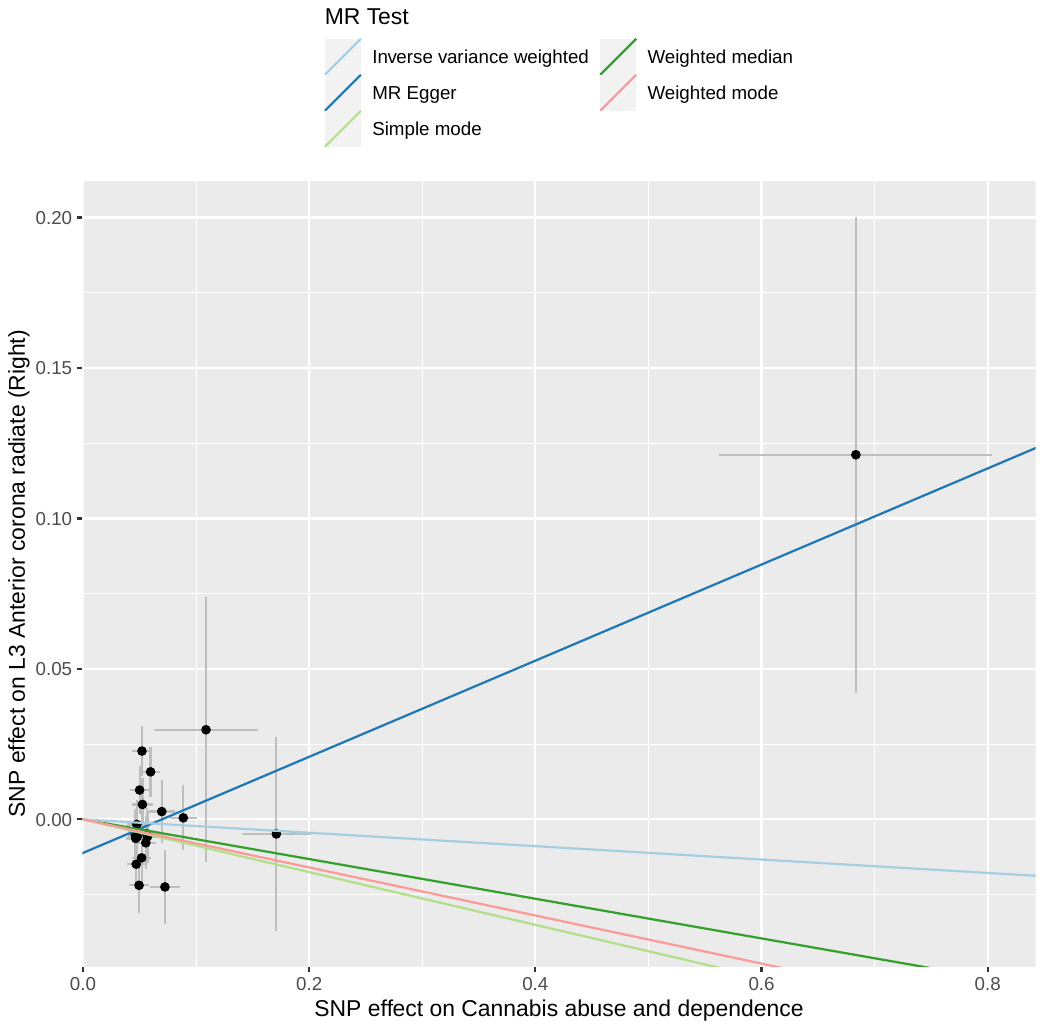

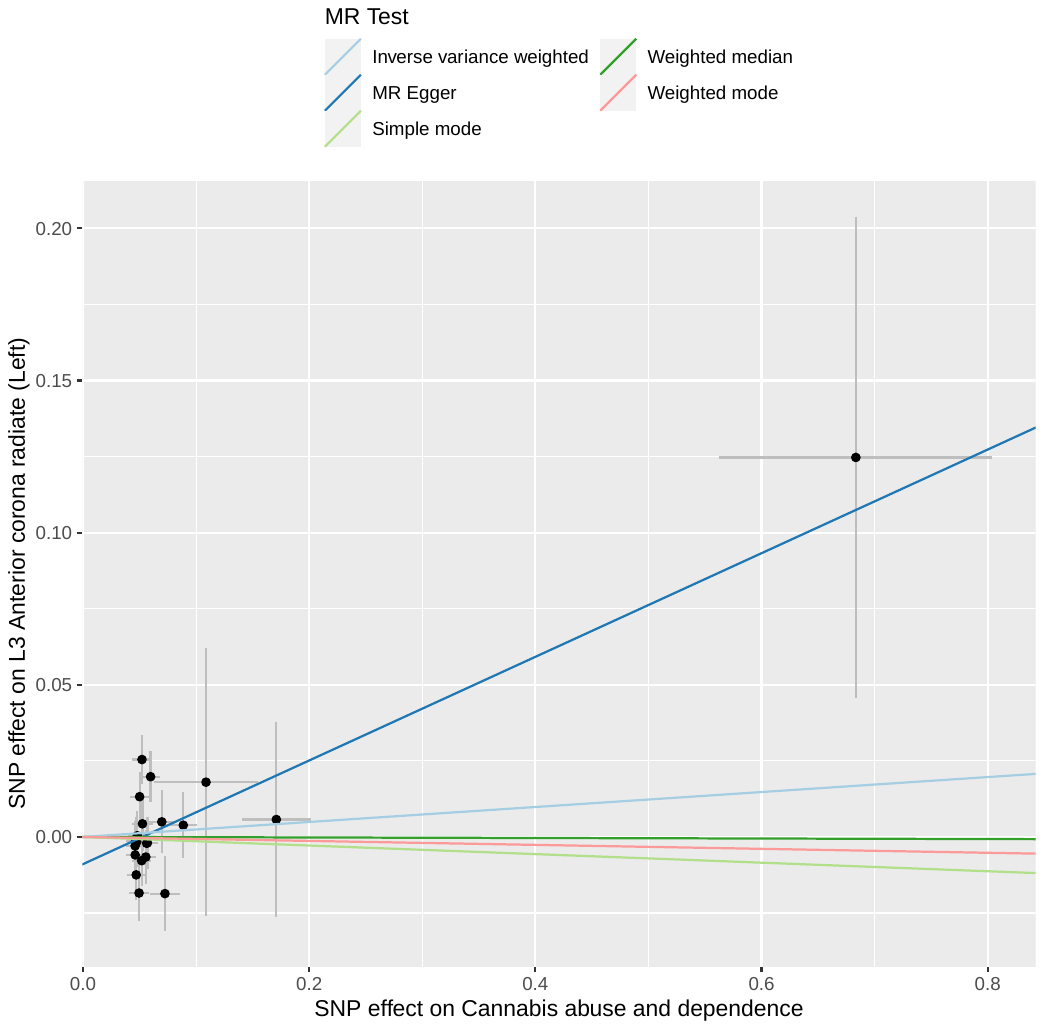

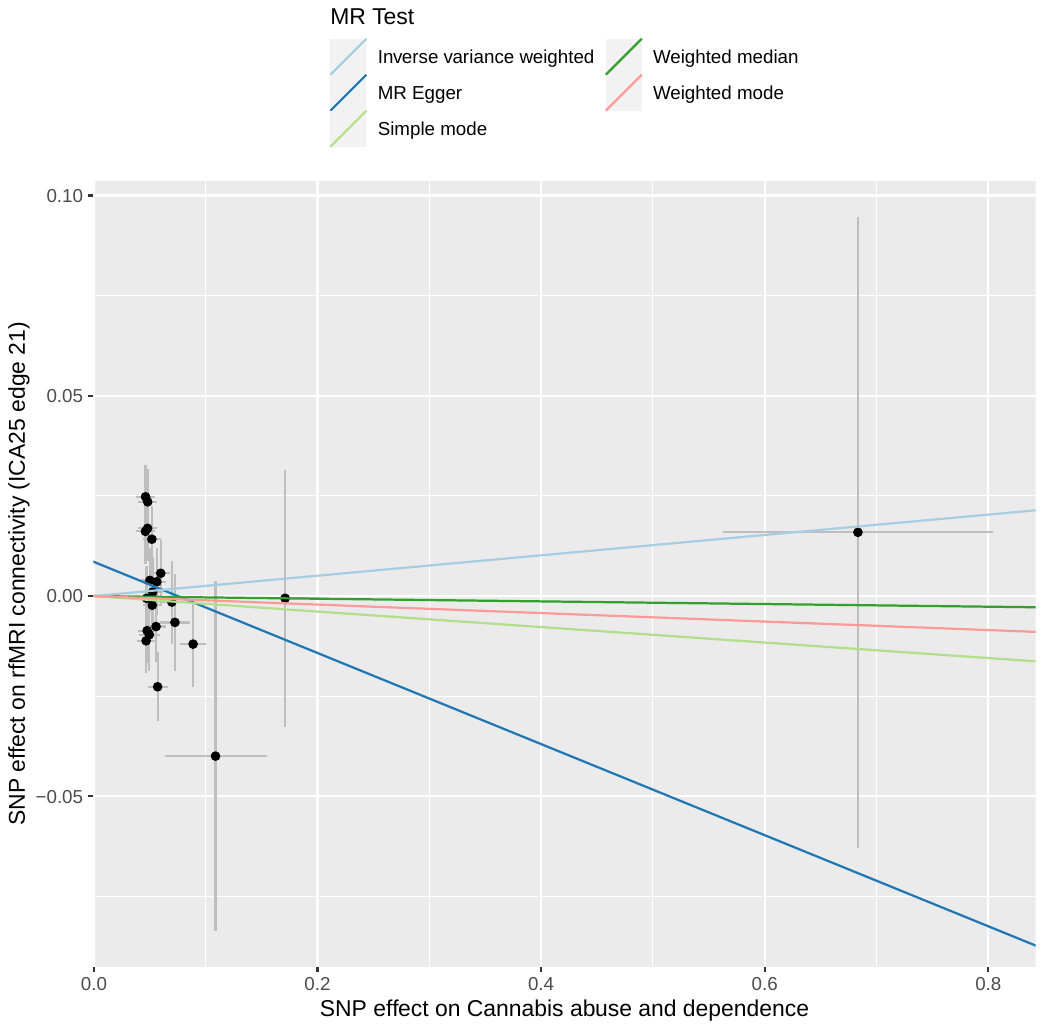

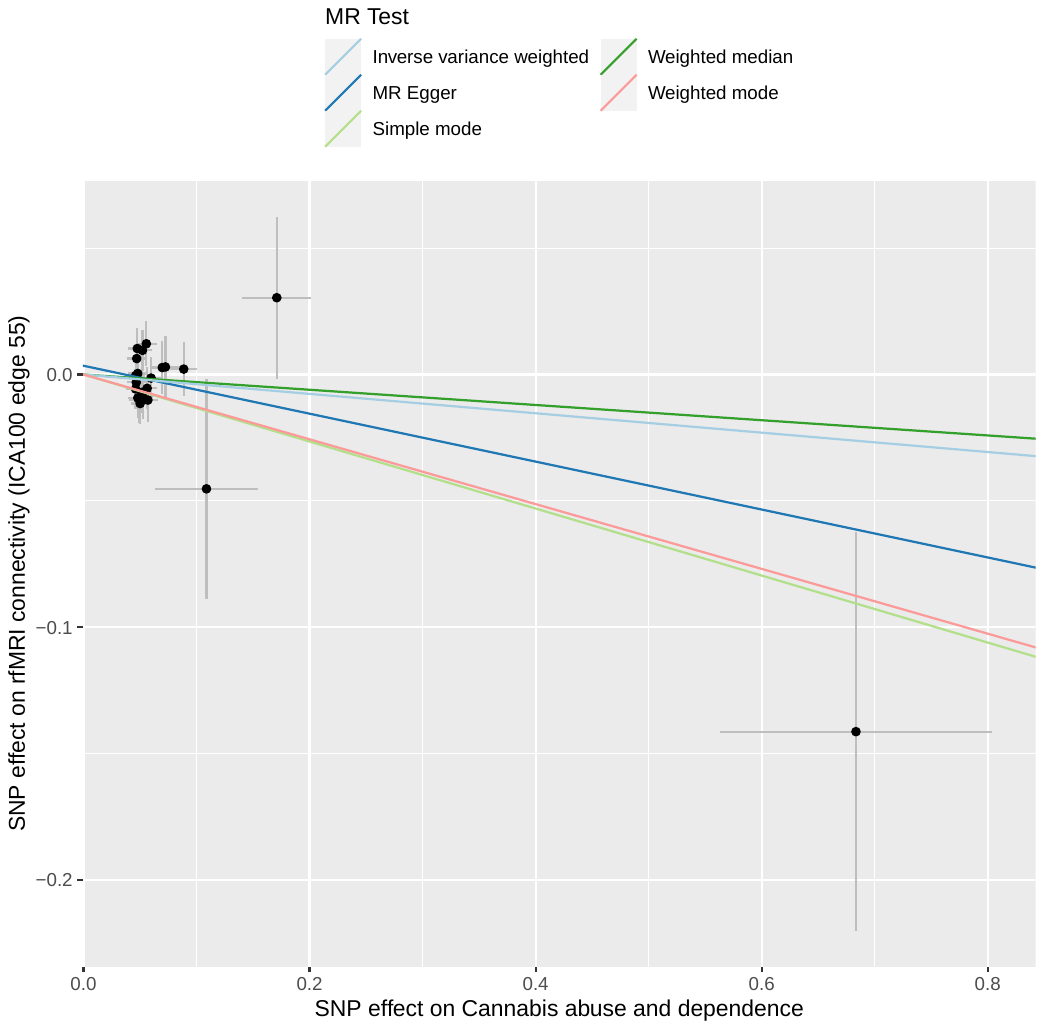

**Two-sample linear MR plot for the causal effect of Lifetime Cannabis use on brain IDPs**

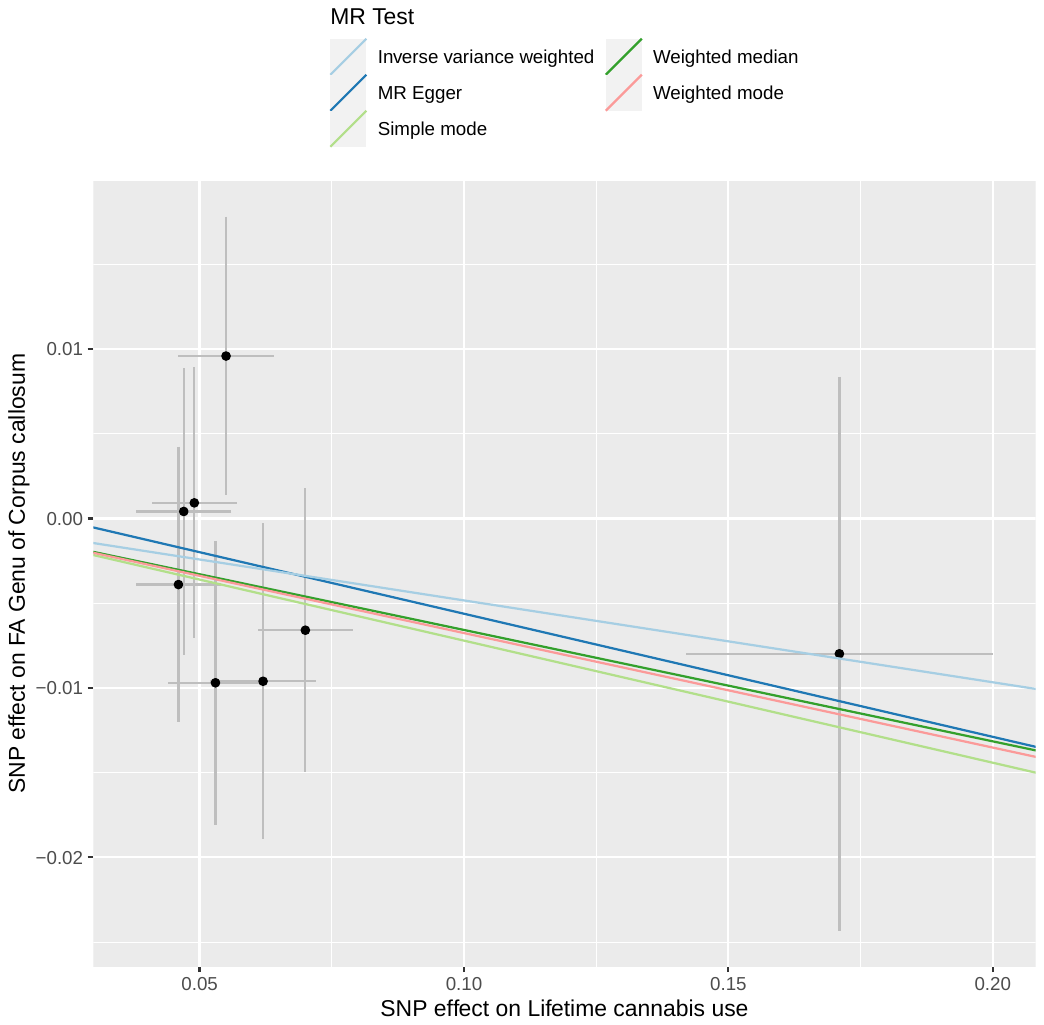

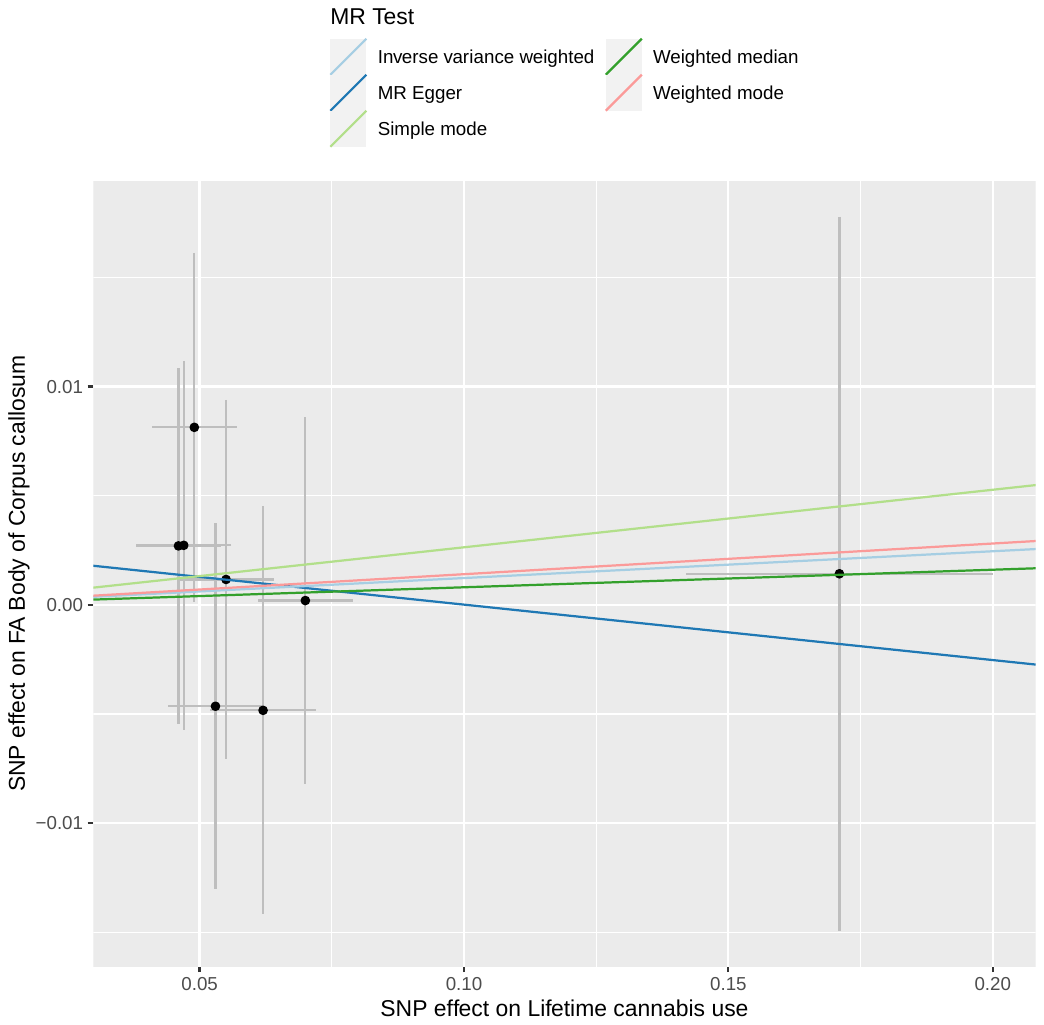

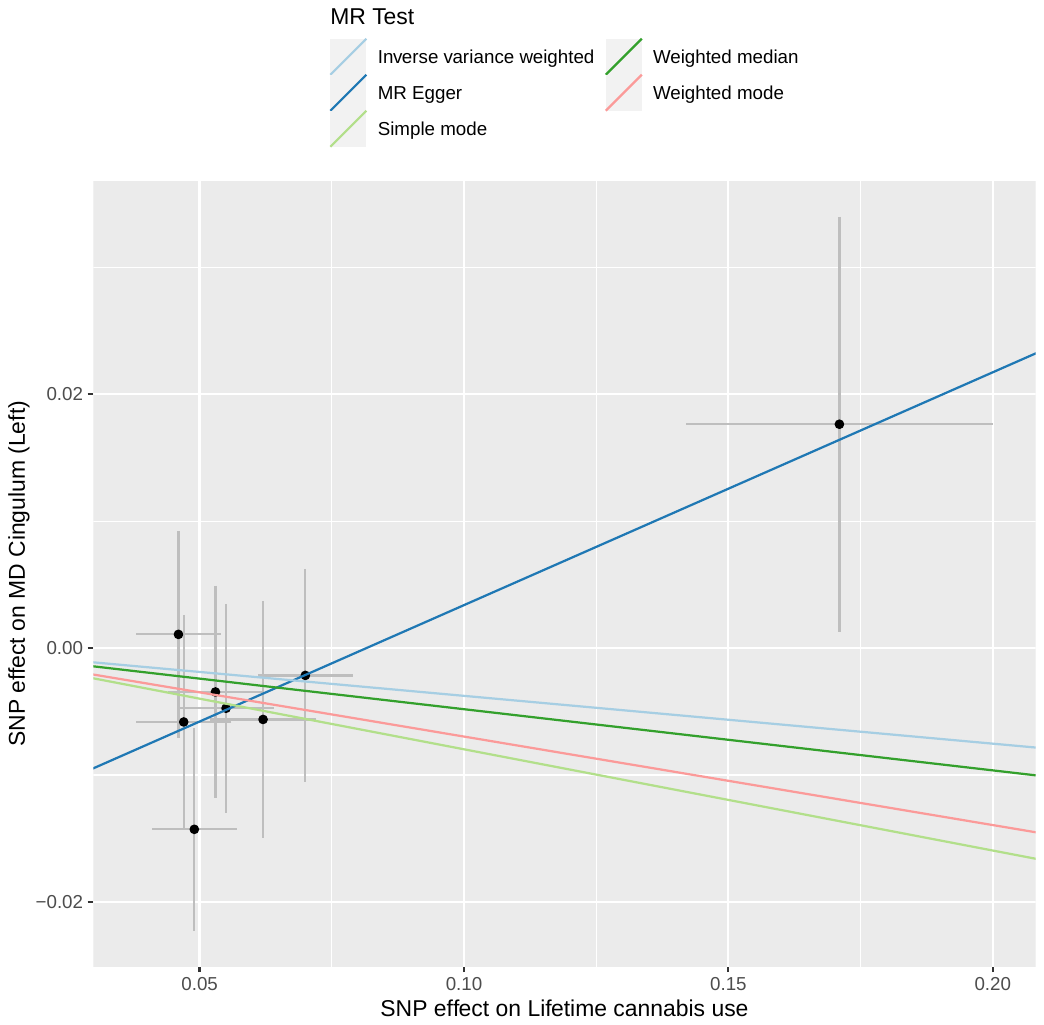

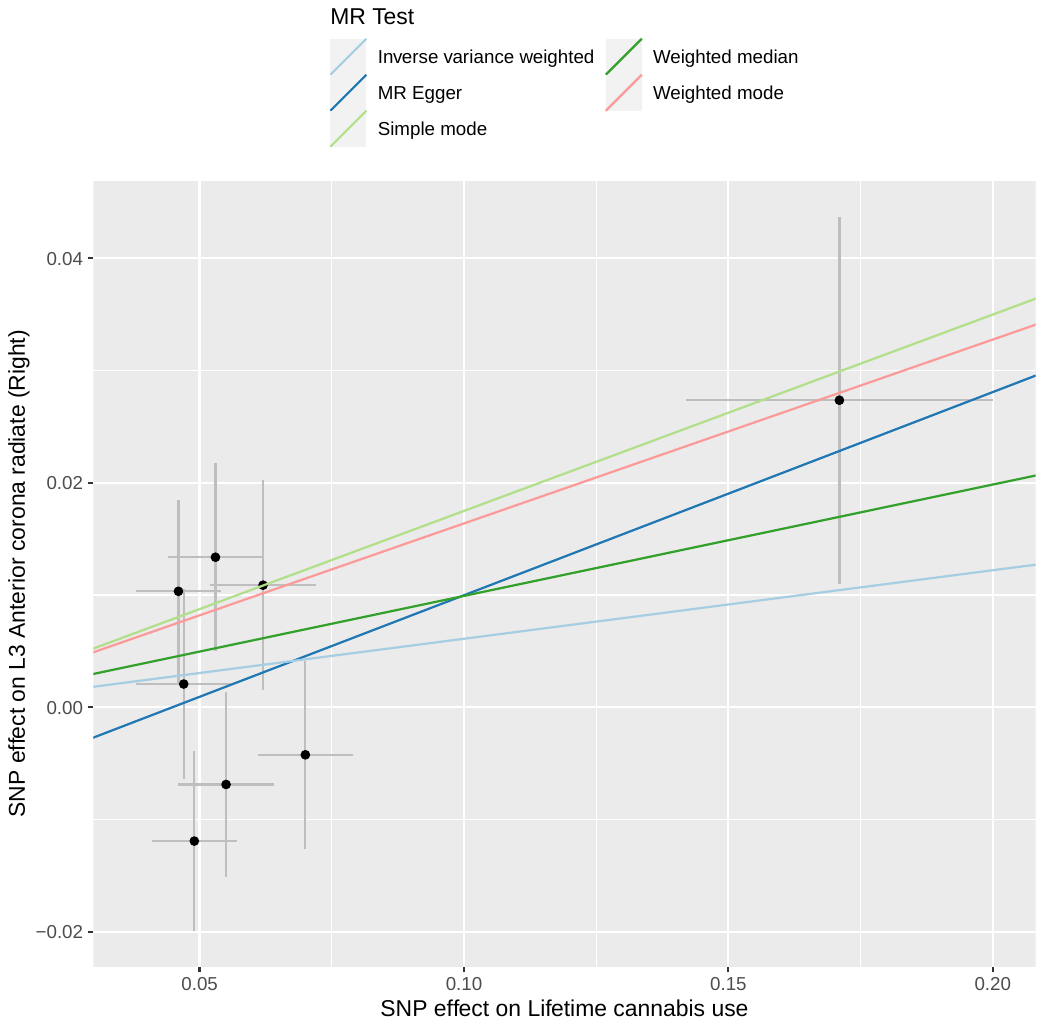

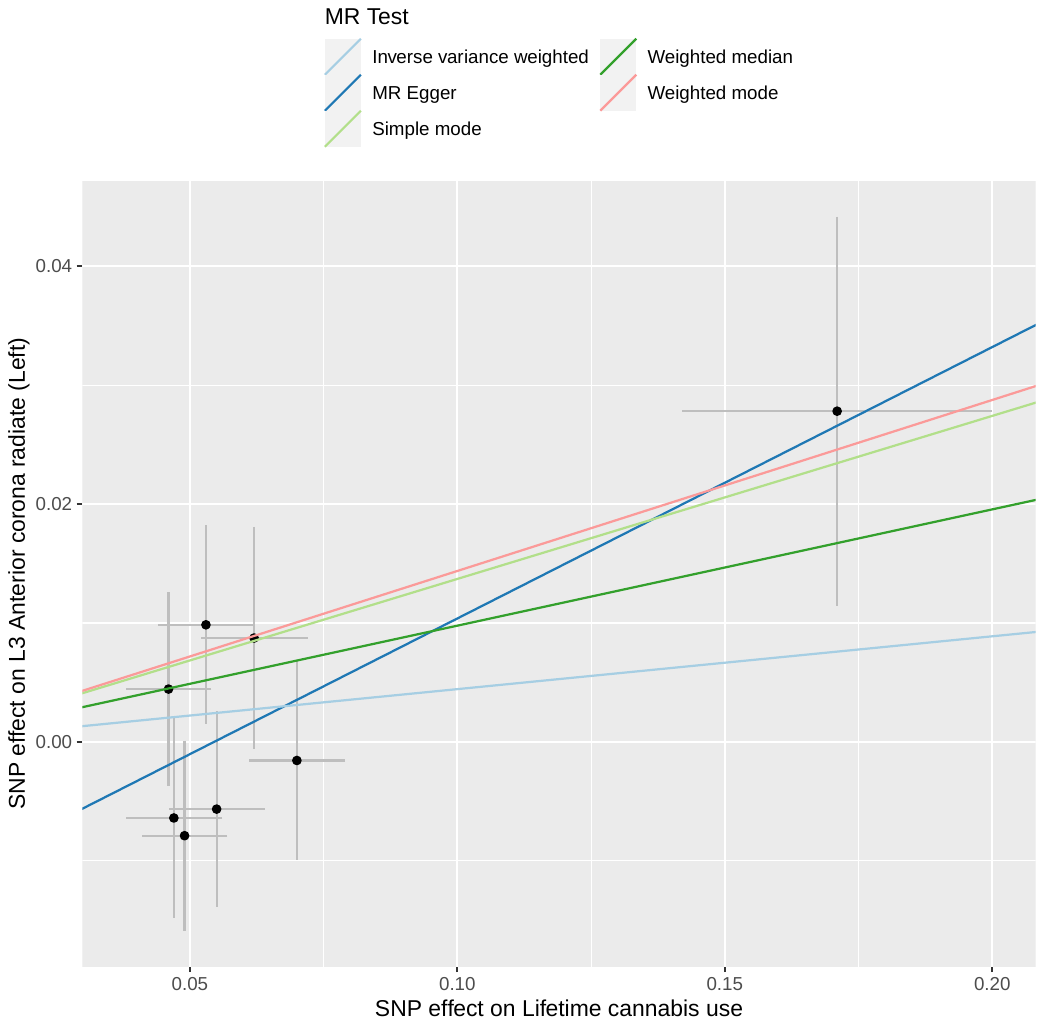

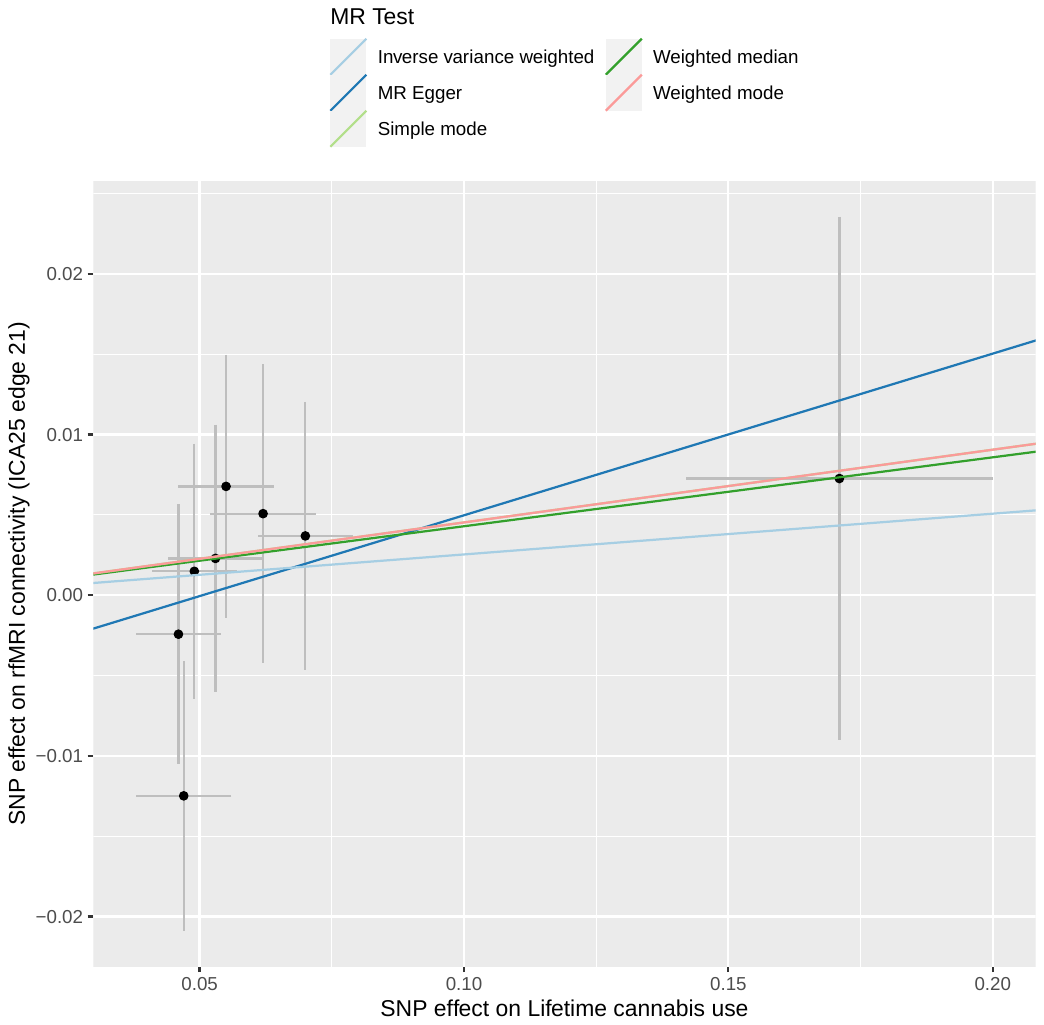

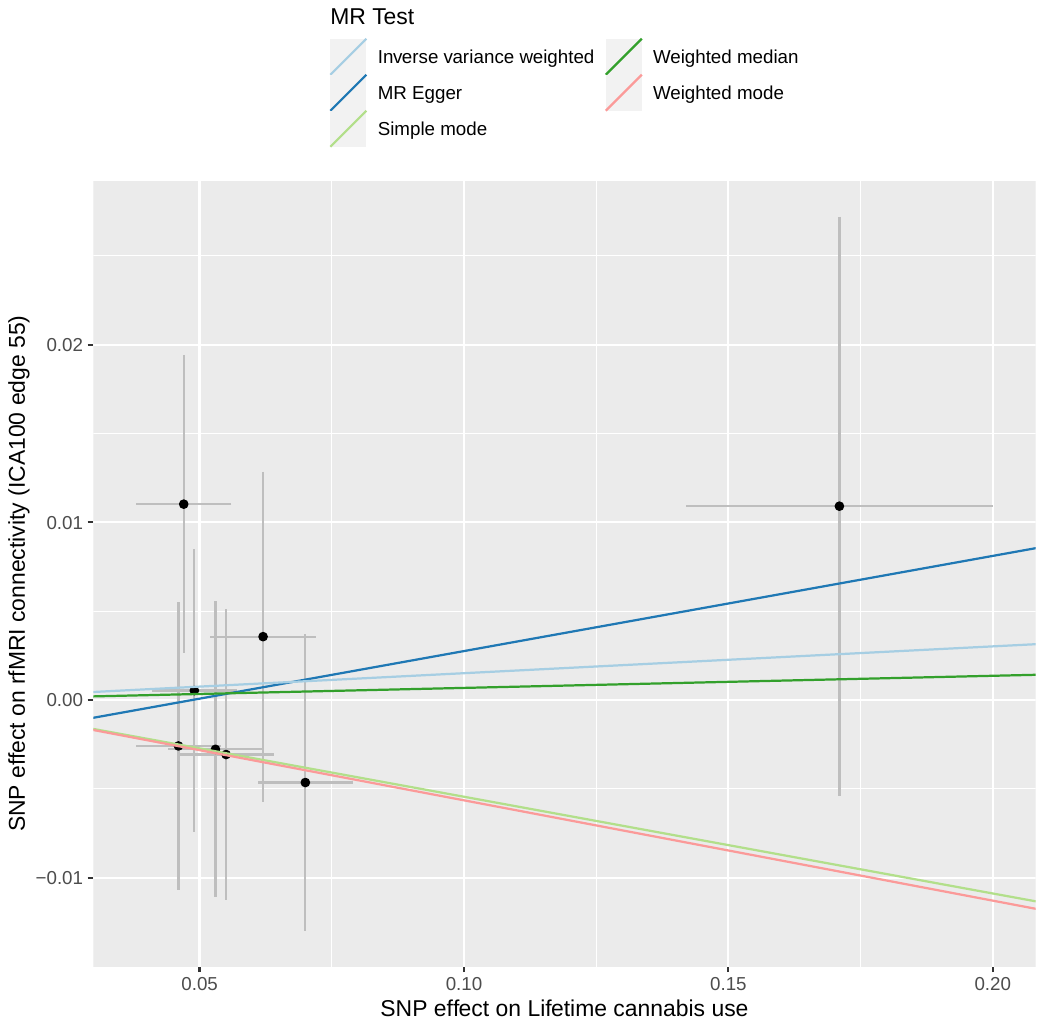

**SFigure 3 (b)**

**Reverse two-sample linear MR plot for the causal effect of brain IDPs on Cannabis dependence or abuse**

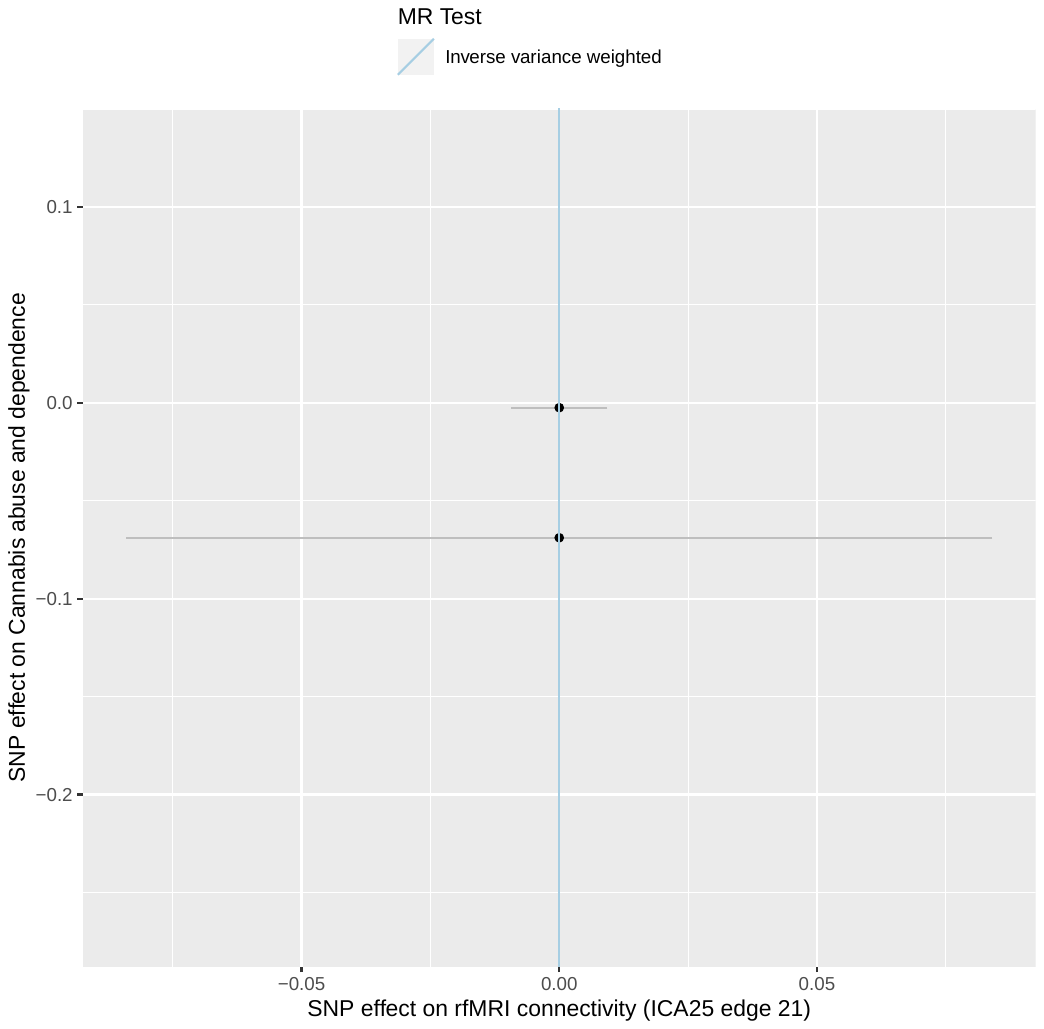

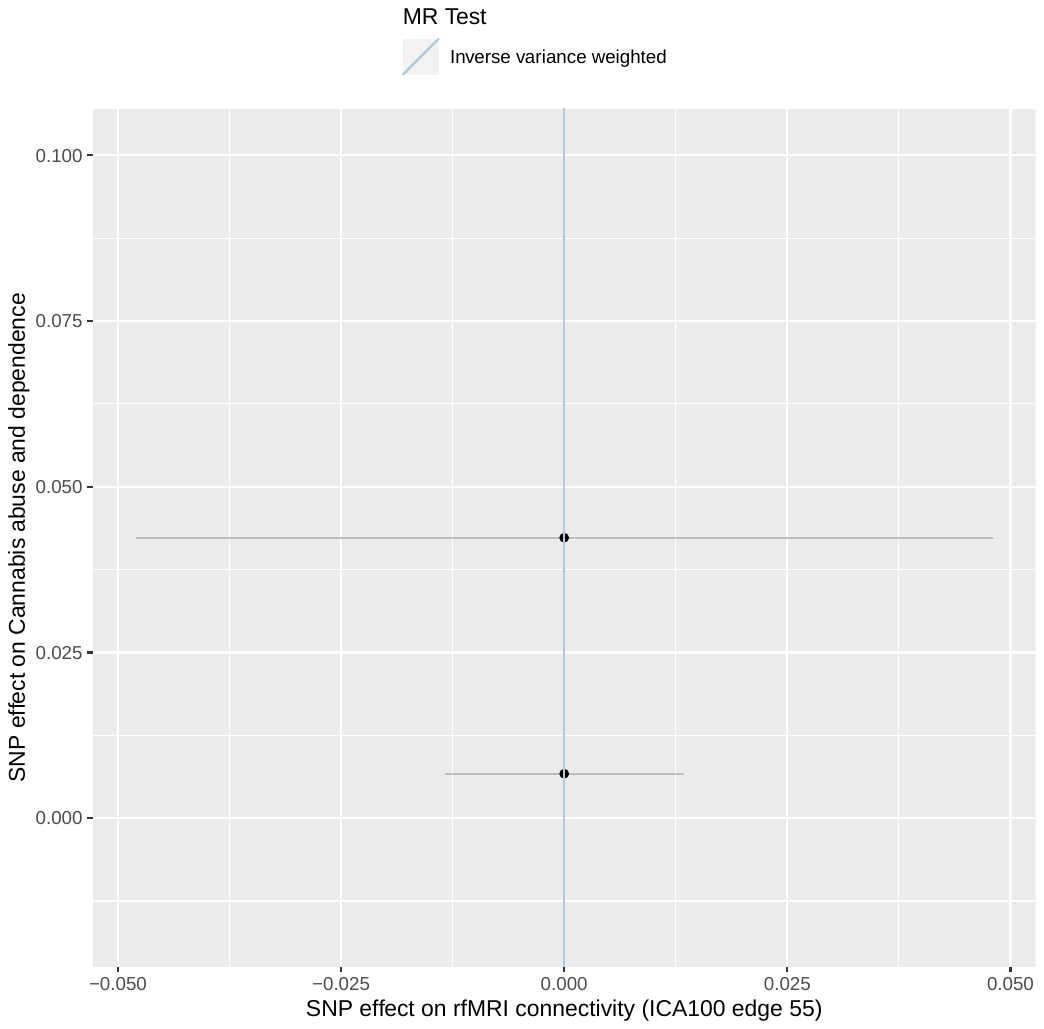
