## Supplemental Table 1 for "Association between cannabis use and brain imaging phenotypes in UK Biobank: an observational and Mendelian randomization study"

**STable 1 (a): SNPs identified in GWAS associated with cannabis use**

| **GWAS** | **SNP** | **Chr** | **BP** | **A1** | **A2** | **β** | **SE** | ***P*-value** |
| --- | --- | --- | --- | --- | --- | --- | --- | --- |
| Cannabis dependence or abuse  (Levey et al 2023) | rs10835372 | 11 | 28643913 | C | T | 0.047 | 0.0085 | 3.028e-08 |
|  | rs10986600 | 9 | 127928735 | C | T | 0.057 | 0.0090 | 2.172e-10 |
|  | rs11711407 | 3 | 50225029 | G | A | 0.046 | 0.008 | 2.954e-08 |
|  | rs1526480 | 1 | 91209986 | C | T | -0.052 | 0.008 | 5.906e-10 |
|  | rs159365 | 5 | 60500273 | G | A | 0.046 | 0.008 | 3.330e-08 |
|  | rs17007864 | 3 | 70876858 | C | T | 0.052 | 0.009 | 1.053e-09 |
|  | rs62461183 | 7 | 77716309 | C | T | 0.07 | 0.011 | 5.863e-10 |
|  | rs73247642* | 4 | 47114175 | C | T | -0.109 | 0.046 | 1.699e-02 |
|  | rs11608109* | 11 | 113303448 | C | G | -0.048 | 0.008 | 1.550e-08 |
|  | rs1637570 | 10 | 118619529 | A | G | -0.05 | 0.009 | 4.786e-08 |
|  | rs2014920 | 11 | 113466565 | T | G | 0.053 | 0.009 | 8.004e-09 |
|  | rs2189010 | 7 | 114119430 | A | G | 0.048 | 0.009 | 1.280e-08 |
|  | rs3774800 | 3 | 49334768 | A | G | -0.06 | 0.009 | 1.718e-12 |
|  | rs545943750 | 8 | 16059558 | A | AT | -0.683 | 0.121 | 1.449e-08 |
|  | rs56070621 | 5 | 30825684 | A | T | 0.048 | 0.008 | 1.151e-08 |
|  | rs56372821 | 8 | 27436500 | A | G | -0.089 | 0.012 | 7.272e-14 |
|  | rs62051488 | 16 | 72652784 | A | C | -0.073 | 0.013 | 2.976e-08 |
|  | rs6690119 | 1 | 73580964 | T | C | 0.047 | 0.009 | 4.995e-08 |
|  | rs726610 | 3 | 85551403 | T | C | -0.056 | 0.009 | 4.288e-11 |
|  | rs7519259 | 1 | 66434743 | A | G | 0.05 | 0.008 | 1.830e-09 |
|  | rs80030908 | 13 | 55159898 | A | G | 0.171 | 0.031 | 2.127e-08 |
|  | rs9344740 | 6 | 88619412 | T | G | -0.056 | 0.009 | 8.344e-10 |
| Lifetime cannabis use (Pasman et al 2018) | rs2875907 | 3 | 85518580 | A | G | 0.07 | 0.009 | 9.38E-17 |
|  | rs1448602 | 3 | 85780454 | A | G | -0.062 | 0.01 | 6.55E-11 |
|  | rs7651996 | 3 | 85057349 | T | G | 0.049 | 0.008 | 2.37E-09 |
|  | rs10085617 | 7 | 3634711 | A | T | 0.046 | 0.008 | 2.93E-08 |
|  | rs9773390 | 8 | 81565692 | T | C | -0.171 | 0.029 | 5.66E-09 |
|  | rs9919557 | 11 | 112877408 | T | C | -0.055 | 0.009 | 9.94E-11 |
|  | rs10499 | 16 | 28915527 | A | G | 0.053 | 0.009 | 1.13E-09 |
|  | rs17761723 | 17 | 2107090 | T | C | 0.047 | 0.009 | 3.24E-08 |

Significance threshold was set at *p*<5E-08

*Proxy SNPs

Abbreviations: Chromosome (Chr), location in base pairs (BP), effect allele (A1), allele 2 (A2), Frequency of allele 1 (Freq A1), effect size beta (β), standard error of beta (SE)

**STable 1 (b): SNPs identified in GWAS associated with brain IDPs**

| **GWAS** | **SNP** | **Chr** | **BP** | **A1** | **A2** | **β** | **SE** | ***P*-value** |
| --- | --- | --- | --- | --- | --- | --- | --- | --- |
| FA Genu of Corpus callosum | rs72776055 | 5 | 85744869 | G | T | 3.33e-10 | 0.08 | 1.44e-9 |
| rfMRI connectivity (ICA25 edge 21) | rs559370521 | 5 | 172852051 | T | C | -7.55e-9 | 0.084 | 3.11e-8 |
|  | rs7113557 | 11 | 48317492 | C | T | -5.86e-11 | 0.009 | 2.18e-9 |
| rfMRI connectivity (ICA100 edge 55) | rs147270121 | 5 | 153248944 | C | T | -3.43e-9 | 0.048 | 2.48e-8 |
|  | rs28649975 | 8 | 92275269 | C | T | -1.21e-9 | 0.013 | 3.12e-8 |

Significance threshold was set at *p*<5E-08

Abbreviations: Chromosome (Chr), location in base pairs (BP), effect allele (A1), allele 2 (A2), Frequency of allele 1, effect size beta (β), standard error of beta (SE)
