## Supplemental Table 2 for "Association between cannabis use and brain imaging phenotypes in UK Biobank: an observational and Mendelian randomization study"

**STable 2: Association between cannabis use with grey matter measures.**

|  | **All/Lifetime cannabis use vs. controls** | | | |
| --- | --- | --- | --- | --- |
|  | **Beta** | **LCI** | **UCI** | **P-value uncorrected** |
| Hippocampus Left | 0.026 | -0.013 | 0.062 | 0.183 |
| Hippocampus Right | 0.010 | -0.028 | 0.045 | 0.607 |
| Amygdala Left | 0.036 | -0.005 | 0.072 | 0.073 |
| Amygdala Right | 0.031 | -0.01 | 0.069 | 0.122 |

Estimates were generated using multiple linear regression models adjusted for: age, sex, Townsend deprivation index, employment status, educational qualifications, alcohol drinking status, smoking status, body mass index, systolic and diastolic blood pressure, assessment center, nerves/anxiety/tension/depression status and brain imaging confounds.

Abbreviations: LCI, lower confidence interval; UCI, upper confidence interval
