## Supplemental Table 5 for "Association between cannabis use and brain imaging phenotypes in UK Biobank: an observational and Mendelian randomization study"

**STable 5 (a): Two-sample linear MR estimates for the causal effect of cannabis use on brain IDPs**

| **Brain IDPs** | **SNPs** | **Estimates** | **SE** | ***P*-value** |
| --- | --- | --- | --- | --- |
| FA Genu of Corpus callosum | 22* | 0.01 | 0.053 | 0.849 |
|  | 8** | -0.048 | 0.049 | 0.329 |
| FA Body of Corpus callosum | 22* | 0.04 | 0.042 | 0.333 |
|  | 8** | 0.012 | 0.049 | 0.804 |
| MD Cingulum cingulate gyrus (Left) | 22* | 0.032 | 0.039 | 0.405 |
|  | 8** | -0.038 | 0.049 | 0.447 |
| L3 Anterior corona radiate (Right) | 22* | -0.022 | 0.044 | 0.61 |
|  | 8** | 0.061 | 0.059 | 0.303 |
| L3 Anterior corona radiate (Left) | 22* | 0.025 | 0.042 | 0.559 |
|  | 8** | 0.044 | 0.049 | 0.369 |
| rfMRI connectivity (ICA25 edge 21) | 22* | 0.025 | 0.049 | 0.606 |
|  | 8** | 0.025 | 0.049 | 0.607 |
| rfMRI connectivity (ICA100 edge 55) | 22* | -0.038 | 0.034 | 0.26 |
|  | 8** | 0.015 | 0.049 | 0.759 |

Estimates are from inverse-variance weighted (IVW) analysis.

* SNPs associated with *cannabis dependence and abuse*

** SNPs associated *with lifetime cannabis use*

Abbreviations: IDP, imaging-derived phenotype; SNP, single nucleotide polymorphism; SE, standard error.

**STable 5 (b): Two-sample linear Reverse MR estimates for the causal effect of brain IDPs on cannabis use**

| **Cannabis use** | **SNPs** | **Estimates** | **SE** | ***P*-value** |
| --- | --- | --- | --- | --- |
| Cannabis dependence or abuse | 1* | -283^a^ | 266 | 0.915 |
|  | 2** | -988^b^ | 261 | 0.706 |
|  | 2*** | -767^b^ | 956 | 0.423 |
| Lifetime cannabis use | 1* | -1837.928^a^ | 885.547 | 0.038 |
|  | 1* | 20.273^a^ | 40.283 | 0.615 |

^a^ Estimates are from Wald ration

^b^ Estimates are from Inverse-variance weighted (IVW) analysis

* SNPs associated with FA genu of corpus callosum

** SNPs associated with rfMRI connectivity (ICA25 edge 21)

** SNPs associated with rfMRI connectivity (ICA100 edge 55)

Abbreviations: IDP, imaging-derived phenotype; SNP, single nucleotide polymorphism; SE, standard error.
